# Cohort Design and Natural Language Processing to Reduce Bias in Electronic Health Records Research: The Community Care Cohort Project

**DOI:** 10.1101/2021.05.26.21257872

**Authors:** Shaan Khurshid, Christopher Reeder, Lia X. Harrington, Pulkit Singh, Gopal Sarma, Samuel F. Friedman, Paolo Di Achille, Nathaniel Diamant, Jonathan W. Cunningham, Ashby C. Turner, Emily S. Lau, Julian S. Haimovich, Mostafa A. Al-Alusi, Xin Wang, Marcus D.R. Klarqvist, Jeffrey M. Ashburner, Christian Diedrich, Mercedeh Ghadessi, Johanna Mielke, Hanna M. Eilken, Alice McElhinney, Andrea Derix, Steven J. Atlas, Patrick T. Ellinor, Anthony A. Philippakis, Christopher D. Anderson, Jennifer E. Ho, Puneet Batra, Steven A. Lubitz

## Abstract

**Background:** Electronic health records (EHRs) promise to enable broad-ranging discovery with power exceeding that of conventional research cohort studies. However, research using EHR datasets may be subject to selection bias, which can be compounded by missing data, limiting the generalizability of derived insights.

**Methods:** Mass General Brigham (MGB) is a large New England-based healthcare network comprising seven tertiary care and community hospitals with associated outpatient practices. Within an MGB-based EHR warehouse of >3.5 million individuals with at least one ambulatory care visit, we approximated a community-based cohort study by selectively sampling individuals longitudinally attending primary care practices between 2001-2018 (n=520,868), which we named the Community Care Cohort Project (C3PO). We also utilized pre-trained deep natural language processing (NLP) models to recover vital signs (i.e., height, weight, and blood pressure) from unstructured notes in the EHR. We assessed the validity of C3PO by deploying established risk models including the Pooled Cohort Equations (PCE) and the Cohorts for Aging and Genomic Epidemiology Atrial Fibrillation (CHARGE-AF) score, and compared model performance in C3PO to that observed within typical EHR Convenience Samples which included all individuals from the same parent EHR with sufficient data to calculate each score but without a requirement for longitudinal primary care. All analyses were facilitated by the JEDI Extractive Data Infrastructure pipeline which we designed to efficiently aggregate EHR data within a unified framework conducive to regular updates.

**Results:** C3PO includes 520,868 individuals (mean age 48 years, 61% women, median follow-up 7.2 years, median primary care visits per individual 13). Estimated using reports, C3PO contains over 2.9 million electrocardiograms, 450,000 echocardiograms, 12,000 cardiac magnetic resonance images, and 75 million narrative notes. Using tabular data alone, 286,009 individuals (54.9%) had all vital signs available at baseline, which increased to 358,411 (68.8%) after NLP recovery (31% reduction in missingness). Among individuals with both NLP and tabular data available, NLP-extracted and tabular vital signs obtained on the same day were highly correlated (e.g., Pearson r range 0.95-0.99, p<0.01 for all). Both the PCE models (c-index range 0.724-0.770) and CHARGE-AF (c-index 0.782, 95% 0.777-0.787) demonstrated good discrimination. As compared to the Convenience Samples, AF and MI/stroke incidence rates in C3PO were lower and calibration error was smaller for both PCE (integrated calibration index range 0.012-0.030 vs. 0.028-0.046) and CHARGE-AF (0.028 vs. 0.036).

**Conclusions:** Intentional sampling of individuals receiving regular ambulatory care and use of NLP to recover missing data have the potential to reduce bias in EHR research and maximize generalizability of insights.

## Introduction

Electronic health record (EHRs) databases are increasingly recognized as powerful tools for biological discovery and clinical insight.^1^ EHR databases provide favorable statistical power for large-scale association (e.g., epidemiological, genetic) analyses, rich and diverse feature sets including clinical risk factors, laboratory results, free text notes, and raw imaging data,^2–5^ and repeated measures to support modeling of disease progression and clinical trajectories.^6^ However, EHR data are subject to selection bias introduced by the acquisition of data on the basis of clinical need,^3,7^ and tend to exhibit high rates of missing data.^4,8,9^

Many studies utilize EHR-derived samples including all available individuals for a particular modeling application. Such an approach is practical and powerful, but resulting samples may be substantially biased, and are more likely to be suitable only for the particular scientific question motivating their construction.^10^ In contrast, an EHR dataset sampled *a priori* to include individuals receiving longitudinal primary care more closely mirrors the design of prospective cohort studies and may result in more generalizable insights as well as the flexibility to support a more diverse array of analytic frameworks.^11,12^ Furthermore, acquisition of all unstructured data, such as free text notes, may provide an opportunity to recover data missing from structured fields. We hypothesized that careful design and attention to missingness would result in sufficient power, reduced bias, and valid prediction performance compared to a conventional EHR based analysis.

In the current study, we developed an EHR-based cohort within a multi-institutional healthcare system comprising individuals selected on the basis of receiving regular in-network primary care, to enable novel cardiovascular discovery, which we named the Community Care Cohort Project (C3PO). We built a standardized data processing pipeline to harmonize a diverse range of structured and unstructured data. To demonstrate the potential to reduce missingness in C3PO, we deployed a deep natural language processing (NLP) model to recover four vital sign features using unstructured notes. We assessed overall validity of C3PO by deploying two established clinical risk scores, and comparing model performance to that observed in Convenience Samples constructed from the same parent EHR but including all individuals with sufficient data to calculate each score (i.e., with no requirement for regular in-network primary care).

## Methods

### Cohort construction

Mass General Brigham (MGB) is a multi-institutional healthcare network with a linked EHR spanning seven tertiary care and community hospitals with associated outpatient practices in the New England region of the United States. Study participants were initially identified using an MGB-based data mart containing tabular EHR data for >3.6 million individuals with at least one ambulatory visit between 2000-2018. Given our intent to identify individuals receiving primary care within the MGB system, we developed, validated, and applied rule-based heuristics to identify primary care office visits using Current Procedural Terminology (CPT) codes (**Supplementary Table 1**) and a manually curated list of 431 primary care clinic locations. To increase the probability that individuals received longitudinal primary care within MGB, we restricted the cohort to individuals with at least one pair of primary care visits occurring between 1-3 years apart. To allow for ascertainment of baseline clinical factors, we defined the start of follow-up for each individual as the second primary care visit of that individual’s earliest qualifying pair (**Supplementary Figure 1**).^4^ Study protocols complied with the tenets of the Declaration of Helsinki and were approved by the Mass General Brigham Institutional Review Board.

### Cohort validation

We validated the construction of C3PO using two methods (**Supplementary Methods 1**). First, we assessed overlap between individuals selected for C3PO and an existing sample from a curated Massachusetts General Hospital (MGH) primary care practice registry comprising 297,718 individuals. Prior to applying any temporal and age selection criteria (see below), 93.3% of the MGH registry was included within the candidate set for C3PO. After applying temporal and age selection criteria to the C3PO set, 73.7% of the MGH registry remained included in C3PO (**Supplementary Figure 2**).

Second, we performed manual validation of the EHR for C3PO candidates. Two clinical adjudicators (S.K., M.A.A.) reviewed n=400 records to assess accuracy of the applied selection criteria. Definitions used to determine inclusion and exclusion in C3PO met the pre-specified threshold of ≥85% positive predictive value (PPV) sufficient to proceed with cohort construction (PPV 94% for cases; 92% for non-cases, **Supplementary Table 2**).

### Data ingestion pipeline

After identifying a candidate set of 523,445 individuals in C3PO, we obtained a comprehensive range of EHR data including demographics, anthropometrics and vital signs, narrative notes, laboratory results, medication lists, and radiology/cardiology diagnostic test reports using the Research Patient Data Registry (Boston, Massachusetts), a data repository containing the complete EHR data of all individuals receiving care within MGB.^13^ We then developed a standardized data ingestion pipeline (the JEDI Extractive Data Infrastructure [JEDI]), which integrates a series of distinct files containing an array of different EHR data types into a unified, indexed file system suitable for implementation of an array of traditional and machine learning-based models (Hierarchical Data Format 5^14^). To facilitate interactive data exploration and epidemiologic modeling, we also developed egress pipelines capable of producing customized long-format files (i.e., each row is a distinct observation within the EHR) and wide-format files (i.e., each row is a unique individual and columns represent data from observations, drawing from the full corpus of data available in C3PO). We removed individuals aged <18 or ≥90 years at the start of follow-up, as well as an additional 21 individuals with missing demographic data, resulting in 520,868 individuals in the final C3PO cohort (**Figure 1**).

**Figure 1.**
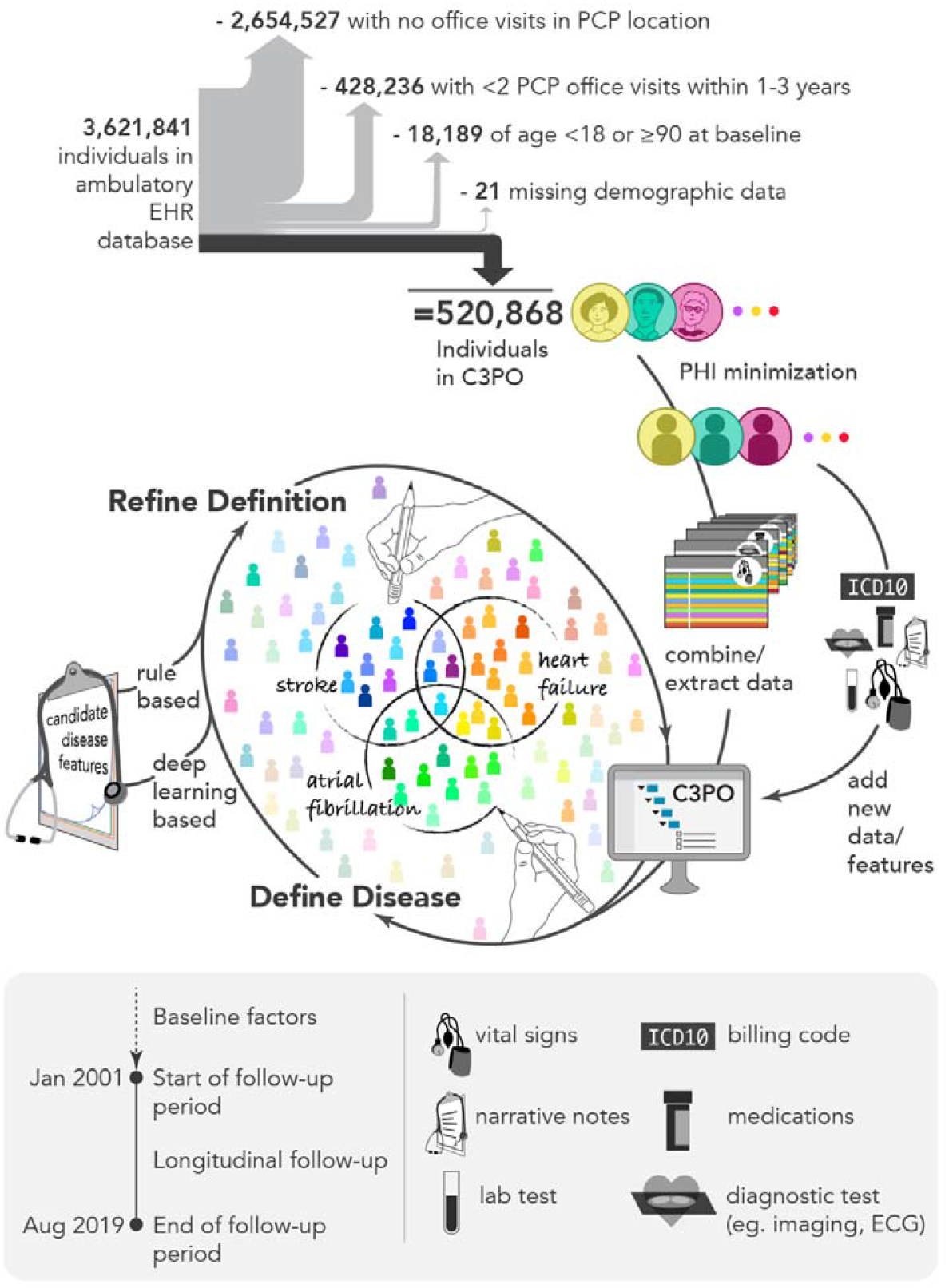
Overview of C3PO construction and data pipeline. Depicted is a graphical overview of the construction of the Community Care Cohort Project (C3PO). C3PO comprises the electronic health record (EHR) data of 520,868 individuals aged 18-90 at the start of sample follow-up, selected from an ambulatory EHR database on the basis of receiving periodic primary care (i.e., ≥2 visits within 1-3 consecutive years, see text). C3PO is structured as an indexed file system containing protected health information-minimized data of various types (bottom panel). The C3PO database can readily accommodate updating of existing data, integration of new data features, and construction of composite disease phenotypes based on multiple data features.

### Cohort validation

We implemented two well-validated and clinically relevant cardiovascular risk prediction models to assess the validity of C3PO as a tool for patient-oriented discovery research and longitudinal risk prediction: 1) the Pooled Cohort Equations (PCE),^15^ and 2) the Cohorts for Aging and Genomic Epidemiology Atrial Fibrillation (CHARGE-AF) score.^16^

Relevant exposures were derived from the EHR. Demographics including age, sex, and race were extracted from EHR demographic data. Height, weight, blood pressure, and smoking status were derived from tabular EHR data extracted from clinical encounters, where the value most closely preceding start of follow-up (within three years) was used, with the exception of height for which any value in the EHR was accepted. Prevalent diseases were defined using previously published groupings of International Classification of Diseases, 9^th^ and 10^th^ revision (ICD-9 and 10) diagnosis codes and CPT codes. A complete list of exposure definitions is shown in **Supplementary Table 3**.

Primary outcomes included the prediction targets for each risk score (i.e., myocardial infarction or ischemic stroke [MI/stroke] for PCE and AF for CHARGE-AF). AF was defined using a previously validated EHR-based AF classification scheme (PPV 92%).^17^ Stroke and myocardial infarction were defined using the presence of ≥2 ICD-9 or ICD-10 codes using previously validated code sets (PPV ≥85%).^18^

### Convenience Samples

We additionally deployed the PCE and CHARGE-AF scores within samples derived from the same parent EHR but constructed solely on the basis of available score components to compare contrasting EHR-based sampling strategies (“PCE Convenience Sample” and “CHARGE-AF Convenience Sample”). Specifically, each Convenience Sample comprised all individuals with each component of the relevant score available within a three-year window (i.e., no requirement for longitudinal primary care). In each Convenience Sample, the start of follow-up began at the earliest time all necessary data became available. Flow diagrams summarizing the construction of the Convenience Samples are shown in **Supplementary Figure 3**. Further details of Convenience Sample construction are described in **Supplementary Methods 2**.

### Natural language processing vital sign recovery

Given relatively high missingness rates for baseline vital signs (>40%), we employed a natural language processing (NLP) algorithm to recover vital signs (i.e., height, weight, systolic and diastolic blood pressures) from unstructured notes. Our NLP methods are described in detail in **Supplementary Methods 3**. Briefly, we utilized Bio+Discharge Summary BERT, a deep contextual word embedding model that has been pretrained consecutively on large corpora of general English text (e.g., Wikipedia), biomedical text (PubMed abstracts and PubMed Central full-text articles),^19^ and physician-written Discharge Summaries (from the MIMIC-III v1.4 database).^20,21^ Such domain-specific pretraining has been shown to yield performance improvements on clinical NLP tasks like named entity recognition, which is relevant for feature recovery.^19,20^ To recover vital signs, we created a rule-based approach to automatically label the position of relevant features and their corresponding units in several different types of clinical notes (e.g., discharge summaries, outpatient progress notes) (**Supplementary Table 4**). We fine-tuned Bio+Discharge SummaryBERT on approximately 120,000 instances of vital signs identified with the rule-based approach. We then imposed physiological constraints and performed unit harmonization on NLP extracted values. Fifty random values of each vital sign type extracted from a held out validation set were inspected by a study cardiologist and had a PPV of 100% for representing the true vital sign of interest. We ran inference with this model on 9,522,262 notes for the 401,826 patients who had eligible notes in the three years prior to start of cohort follow-up, and utilized NLP recovered values in our prediction models for individuals in whom baseline values were missing in the tabular data.

### Statistical analysis

We tabulated the number of cardiac imaging studies, cardiac diagnostic tests, and unstructured text notes available within the dataset. We also cross-referenced the number of individuals in C3PO in whom genetic data are available for analysis through simultaneous participation in the MGB Biobank biorepository. We compared sets of values obtained from both sources on the same day within 3 years of the start of follow-up to assess agreement between vital signs obtained using tabular data versus NLP. We plotted the distribution of paired values, calculated Pearson correlations, and assessed agreement using Bland-Altman plots. If individuals had multiple same-day value pairs for comparison, only the pair most closely preceding start of follow-up was compared.

We calculated the cumulative incidence of events at their respective time horizons (e.g., 5-year AF for CHARGE-AF, 10-year MI/stroke for PCE) using the Kaplan-Meier method. We also calculated event incidence rates per 1,000 person-years and corresponding confidence intervals using the normal approximation. For all longitudinal analyses, person-time ended at the earliest of an outcome event, death, last encounter of any type in the EHR, age 90, or the administrative censoring date for C3PO (August 31, 2019, **Supplementary Figure 1**).

The linear predictors of the CHARGE-AF^16^ and PCE scores^15^ were calculated using their published coefficients. The analysis set for each score was restricted to individuals without the disease of interest at baseline and within the published age range for each score (i.e., 46-90 years for CHARGE-AF and 40-79 years for PCE). For the purposes of calculating CHARGE-AF, the coefficient associated with White race was attributed to individuals who self-reported as White, but not to individuals of other races.^4,8,22^ Although dedicated PCE models are available only for White and Black individuals, some work suggests that the models developed for White individuals perform better than those developed for Black individuals when deployed within individuals of other races.^23^ Therefore, for the purposes of calculating the PCE, the models developed for Black individuals were utilized for individuals of self-reported Black race, while the models developed for White individuals were utilized for individuals of all other races. We also performed secondary analyses in which the White equations were deployed only among individuals self-identifying as White. Scores were then converted into predicted event probabilities at their respective time horizons using their original published equations.

We assessed model performance by fitting Cox proportional hazards models with the linear predictor of each model as the covariate of interest. We then tabulated the hazard ratio (HR) per standard deviation (SD) increase in score, model discrimination using the inverse probability of censoring weighted c-index,^24^ and model calibration. We assessed model calibration in four ways: 1) visual inspection of predicted versus observed event rates within each decile of predicted risk (with corresponding fitted curves obtained using adaptive hazard regression^25^), 2) performing the Greenwood-Nam-D’Agostino (GND) test, in which a greater chi-squared value and smaller p-value suggest evidence of miscalibration,^26^ 3) calculating the calibration slope, defined as the beta coefficient associated with the linear predictor in a Cox proportional hazards model with the prediction target as the outcome and the linear predictor as the sole covariate and where a value of one indicates optimal calibration,^27^ and 4) quantifying the integrated calibration index (ICI), a measure of the average absolute prediction error weighted by the empirical risk distribution.^25^ We assessed calibration measures for the original models as well as after recalibration to the sample-level baseline hazard to ensure calibration-in-the-large.^27,28^ Confidence intervals for the ICI were obtained using bootstrapping (500-1,000 iterations based on stratum sample size).

We plotted the cumulative risk of AF and MI/stroke according to level of predicted risk using CHARGE-AF and PCE, respectively, to assess the prognostic value of each score. For these analyses, we used thresholds <7.5% vs ≥7.5% for MI/stroke risk (i.e., the threshold used to determine candidacy for statin therapy in current American Heart Association/American College of Cardiology primary prevention guidelines^29^) and <2.5%, ≥2.5-5%, and ≥5% for AF risk (thresholds used in the original CHARGE-AF validation study^16^).

We repeated the analyses described above within the AF and MI/stroke Convenience Samples to compare the results of contrasting EHR sampling approaches. We assessed for differences in model calibration performance in C3PO versus the Convenience Samples by comparing calibration slopes and ICI values using bootstrapping (500-1,000 iterations based on stratum sample size).

### Data availability

MGB source data contain potentially identifying information and cannot be shared publicly. The JEDI data processing pipeline underlying C3PO is currently located in a private GitHub repository (https://github.com/broadinstitute/jedi), to which access will be granted upon request to the corresponding author. JEDI is in the process of being open-sourced under a BSD 3-Clause License and will be made publicly available on GitHub upon completion.

## Results

### C3PO cohort

In total, C3PO comprised 520,868 individuals (mean age 48 years, 61% women) with a median follow-up time of 7.2 years (quartile-1: 2.6, quartile-3: 12.9). Individuals in C3PO had a median of 30 office visits (14, 62), and 13 (6, 26) primary care office visits. By comparison, individuals in the Convenience Samples generally had shorter follow-up and substantially fewer office visits (**Figure 2** and **Supplementary Figure 4**). Detailed characteristics of individuals included in C3PO and each Convenience Sample are shown in **Table 1**. A summary of the diverse array of diagnostic, imaging, narrative note, and genetic data types available for individuals in C3PO is shown in **Supplementary Table 5**.

**Figure 2.**
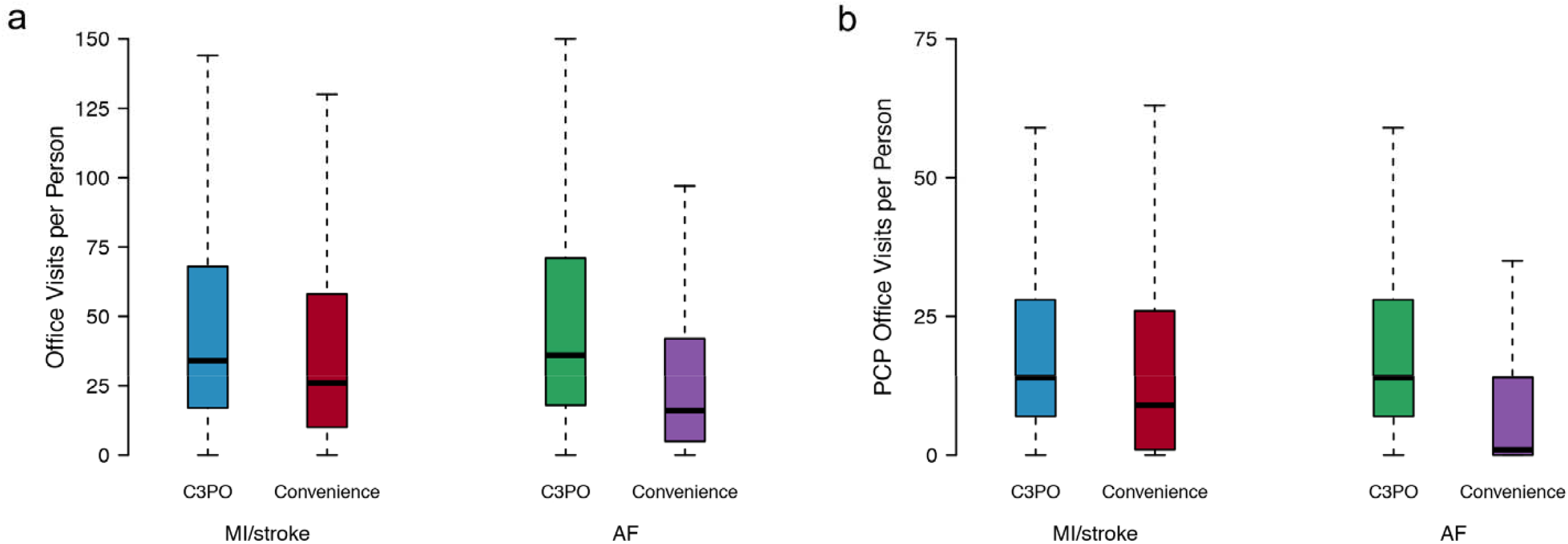
Distribution of office visits in C3PO versus Convenience Samples. Depicted are boxplots demonstrating the distribution of office visits (panel **a**) and primary care physician (PCP) office visits (panel **b**) in the C3PO analysis samples (AF [blue] and MI/stroke [green]) versus the respective Convenience Samples (AF [red] and MI/stroke [purple]). In each boxplot, the black bar denotes the median number of office visits per individual, the box represents the interquartile range, and the whiskers represent points beyond the interquartile range. Points greater than quartile 3 plus 1.5 times the interquartile range and points smaller than quartile 1 minus 1.5 times the interquartile range are not depicted.

**Table 1.** Baseline characteristics

|  | **C3PO\***<br>**(N=520,868)** | **C3PO – MI/stroke**<br>**(N=198,184)<sup>†</sup>** | **MI/stroke**<br>**Convenience**<br>**Sample**<br>**(N=340,226)<sup>†</sup>** | **C3PO – AF**<br>**(N=174,644)<sup>†</sup>** | **AF Convenience**<br>**Sample**<br>**(N=501,272)<sup>†</sup>** |
| --- | --- | --- | --- | --- | --- |
|  | ***Mean ± SD, Median (quartile 1, quartile 3), or N (%)*** |  |  |  |  |
| Age (years) | 48.4 ± 17.1 | 57.0 ± 10.3 | 56.2 ± 10.4 | 60.9 ± 10.0 | 61.4 ± 10.5 |
| Women | 315,577 (60.6%) | 116,448 (58.8%) | 195,039 (57.3%) | 106,279 (60.9%) | 288,334 (57.5%) |
| White | 389,755 (74.8%) | 154,712 (78.1%) | 270,002 (79.4%) | 140,746 (79.6%) | 422,266 (84.2%) |
| Black | 38,104 (7.3%) | 13,805 (7.0%) | 21,248 (6.2%) | 11,103 (6.4%) | 22,787 (4.5%) |
| Hispanic or Latino | 33,762 (6.5%) | 9,401 (4.7%) | 15,142 (4.5%) | 6,804 (3.9%) | 14,115 (2.8%) |
| Asian or Pacific Islander | 21,701 (4.2%) | 7,807 (3.9%) | 13,219 (3.9%) | 6,003 (3.4%) | 14,329 (2.9%) |
| Mixed | 27 (0.05%) | 11 (0.06%) | 24 (0.07%) | 7 (0.04%) | 23 (0.04%) |
| Other | 18,774 (3.6%) | 5,716 (2.9%) | 8,937 (2.6%) | 4,467 (2.6%) | 9,023 (1.8%) |
| Unknown | 18,745 (3.6%) | 6,732 (3.4%) | 11,654 (3.4%) | 5,514 (3.2%) | 18,729 (3.7%) |
| Height (cm) | 167.4 ± 10.4 | - | - | 166.6 ± 10.4 | 167.4 ± 10.3 |
| Weight (kg) | 78.3 ± 20.3 | - | - | 79.4 ± 19.5 | 79.8 ± 19.8 |
| Systolic blood pressure (mmHg) | 123 ± 17 | 126 ± 17 | 127 ± 18 | 128 ± 17 | 130 ± 19 |
| Diastolic blood pressure (mmHg) | 75 ± 10 | - | - | 76 ± 10 | 77 ± 11 |
| Current smoker | 27,202 (5.2%) | 14,720 (7.4%) | 12,652 (3.7%) | 14,031 (8.0%) | 22,020 (4.4%) |
| Anti-hypertensive use | 147,898 (28.4%) | 77,827 (39.3%) | 119,954 (35.3%) | 78,219 (44.8%) | 173,235 (34.6%) |
| Diabetes | 58,159 (11.2%) | 29,307 (14.8%) | 43,966 (12.9%) | 27,953 (16.0%) | 52,180 (10.4%) |
| Heart failure | 12,555 (2.4%) | - | - | 3,334 (1.9%) | 16,786 (3.3%) |
| Myocardial infarction | 17,937 (3.4%) | - | - | 6,641 (3.8%) | 18,260 (3.6%) |
| Total cholesterol (g/dL) | 189 ± 39 | 195 ± 39 | 194 ± 40 | - | - |
| HDL cholesterol (g/dL) | 55 ± 18 | 57 ± 18 | 57 ± 18 | - | - |
| Follow-up, years | 7.2 (2.6, 12.9) | 7.3 (2.8, 11.9) | 7.4 (3.5, 11.8) | 6.5 (2.5, 11.1) | 5.4 (2.2, 9.8) |

\*Values shown exclude missing data
†Only variables relevant for each risk score (CHARGE-AF for AF, PCE for MI/stroke) are depicted

### NLP-based vital sign recovery

Using tabular data alone, 286,009 individuals (54.9%) had height, weight, systolic and diastolic blood pressure available at baseline, which increased to 358,411 (68.8%) after deep learning-enabled NLP recovery (31% reduction in missingness, **Figure 3**). NLP recovery rates stratified by vital sign are shown in **Supplementary Table 6**.

**Figure 3.**
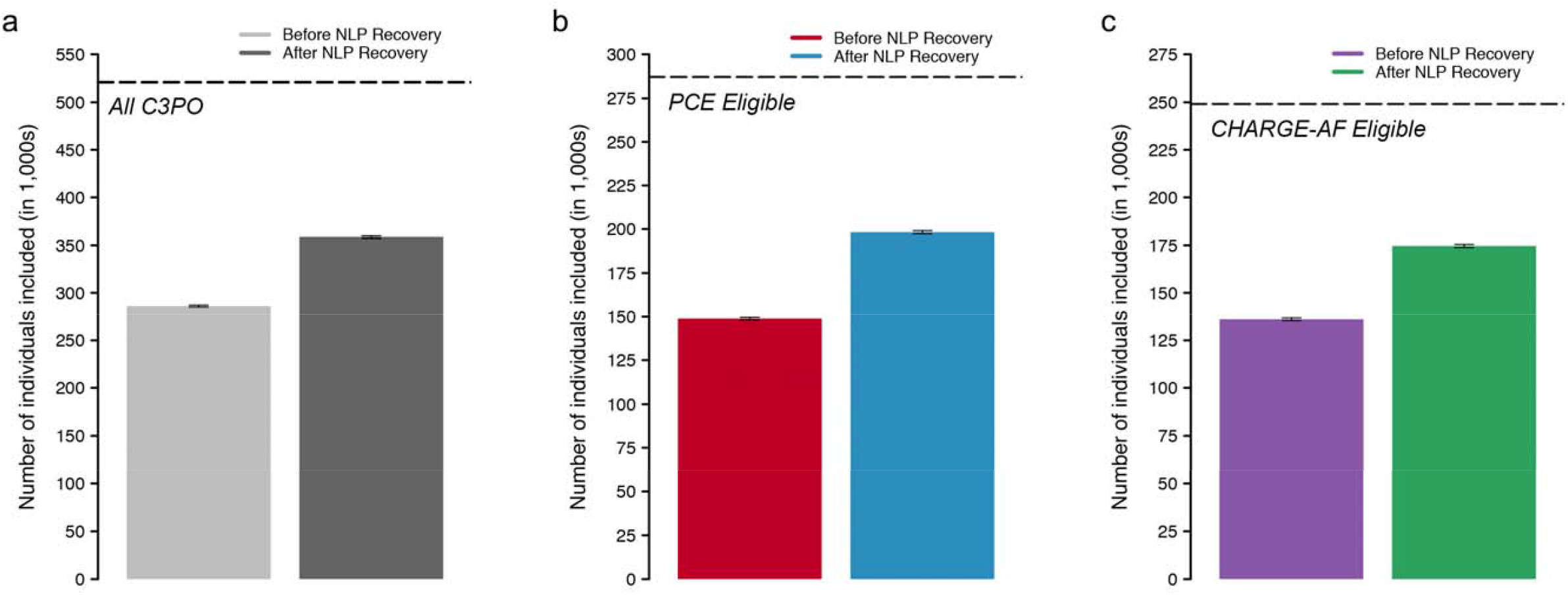
Sample size yield of NLP based missing data recovery. Depicted are relevant sample sizes before and after application of our deep Natural Language Processing (NLP) model to extract missing baseline blood pressure values in C3PO. In panel **a**, the y-axis depicts the total number of individuals with a baseline blood pressure and the hashed line indicates the total sample size of C3PO. In panel **b**, the y-axis depicts the total number of individuals with a complete Pooled Cohort Equations (PCE) score at baseline and the hashed line indicates the total number of individuals eligible for PCE analysis (i.e., within age 40-79 years, with available follow-up data, and without prevalent MI/stroke). In the panel **c**, the y-axis depicts the total number of individuals with a complete CHARGE-AF score at baseline and the hashed line indicates the total number of individuals eligible for CHARGE-AF analysis (i.e., within age 45-94 years, with available follow-up data, and without prevalent AF).

We compared NLP-derived and tabular vital sign data among individuals with values available from both sources on the same day. The distribution of vital sign values was nearly identical using NLP versus tabular data sources (**Figure 4**). The correlation between NLP-derived and tabular vital signs obtained on the same day was excellent (height r=0.99, weight r=0.97, systolic blood pressure r=0.95, diastolic blood pressure r=0.95, p<0.01 for all, **Figure 4**). Intra-individual agreement was generally good (height: -2.97cm to 2.99cm; weight: -8.64kg to 9.29kg; systolic blood pressure: -13.6mmHg to 13.0mmHg; diastolic blood pressure: -8.3mmHg to 8.2mmHg). Bland-Altman plots did not suggest systematic bias (**Figure 4**).

**Figure 4.**
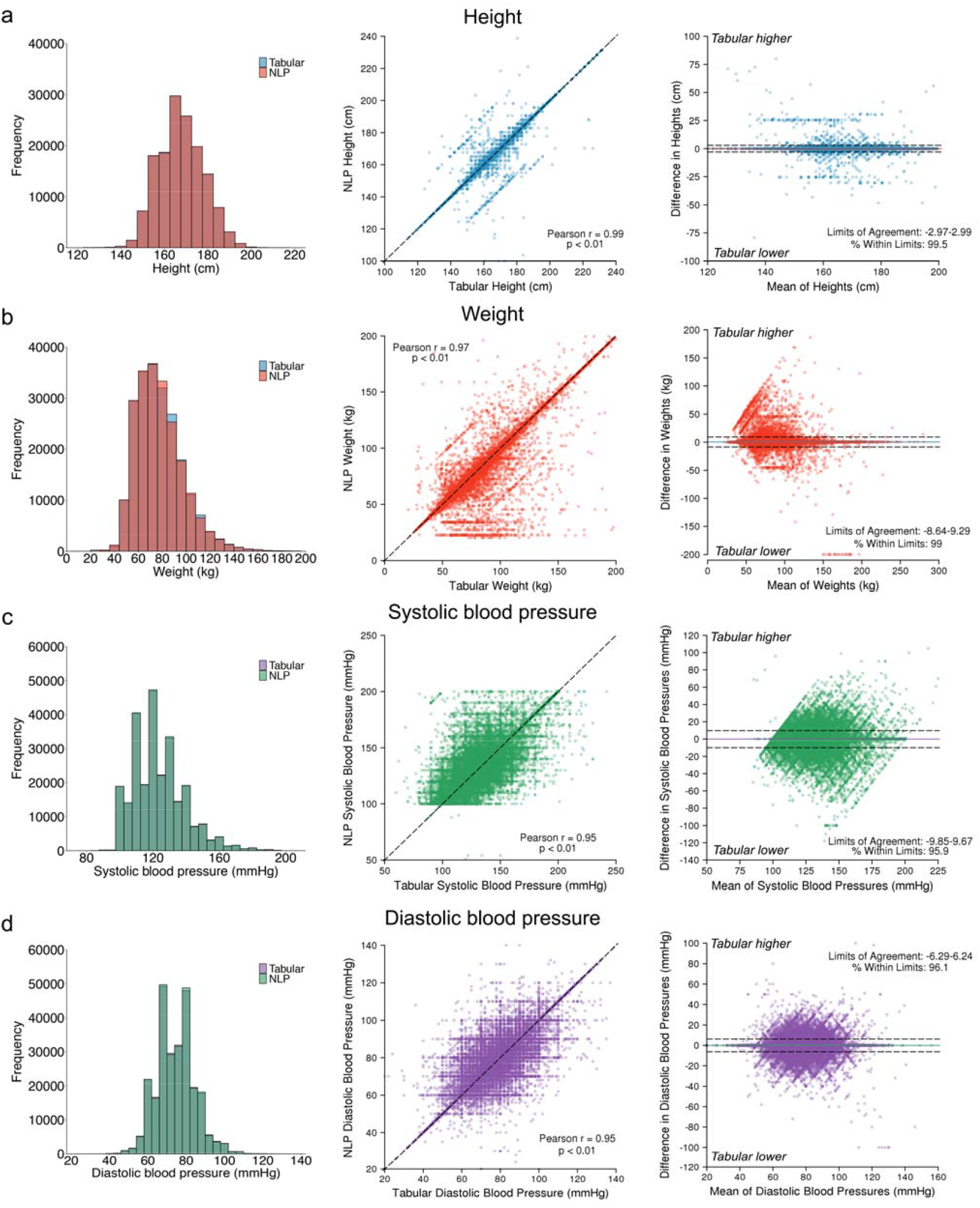
Agreement between tabular and natural language processing-extracted vital signs. Depicted is agreement between vital signs obtained from tabular data and those obtained from our NLP model among individuals with values obtained on the same day. Panels **a** depict height values, panels **b** depict weight values, panels **c** depict systolic blood pressures, and panels **d** depict diastolic blood pressures. For individuals with multiple eligible values, only the pair most closely preceding start of follow-up was used. Left panels show the distribution of values obtained from tabular versus NLP sources. Middle panels show the correlation between tabular values (x-axis) and NLP values (y-axis). Right panels are Bland-Altman plots showing agreement between paired tabular and NLP values. The x-axis depicts increasing mean of the paired values, and the y-axis depicts the difference between the paired values, where positive values denote tabular values greater than corresponding NLP values and negative values denote tabular values lower than corresponding NLP values. The colored horizontal lines depict the mean difference between sources, and the hashed horizontal lines depict 1.96 standard deviations above and below the mean. The values corresponding to the bounds and percentage of values contained within those bounds is printed on each plot.

### MI/stroke analyses – Pooled Cohort Equations

After excluding individuals for missing PCE components, prevalent MI/stroke, absence of follow-up, or age outside 40-79 years, there were a total of 198,184 individuals who were included in incident MI/stroke analyses (**Supplementary Figure 5**). Of the 198,184 individuals in the MI/stroke analysis, 49,289 (24.9%) would have been excluded in the absence of NLP-recovered data (**Figure 3**). At 10 years, there were 10,201 total MI/stroke events (cumulative incidence of MI/stroke 8.0%, 95% CI 7.8-8.1; MI/stroke incidence rate 8.4 per 1,000 person-years, 95% CI 8.2-8.5). The sex-and race-specific PCE scores were each strongly associated with incident MI/stroke (HR per 1-standard deviation (SD) increase range 2.04-2.51), with moderate discrimination (c-index range 0.724-0.768). Miscalibration was present, although relatively modest (GND χ^2^ range 21-487; ICI range 0.012-0.030). Recalibration to the sample average MI/stroke risk did not substantively improve calibration (GND χ^2^ range 18-1,689; ICI range 0.010-0.034; calibration slope range 0.60-0.88). Details of PCE model fit, discrimination, and calibration are shown in **Table 2**. The distribution of predicted MI/stroke risk before and after recalibration is shown in **Supplementary Figure 6**. Cumulative risk of MI/stroke stratified by predicted risk using the original PCE models is shown in **Supplementary Figure 7**. Detailed assessments of PCE calibration before and after recalibration are shown in **Supplementary Figures 8-9**. Results were similar in models deploying the White PCE algorithms only in individuals identifying as White (**Supplementary Table 7**). Model assessment with versus without NLP recovered values demonstrated similar performance but with greater precision in the NLP enabled analyses (**Supplementary Table 8**).

**Table 2.** Risk score performance in C3PO versus Convenience Samples

| Model | Hazard ratio<br>(per 1-SD<br>increase) | C-index<sup>†</sup> | GND $\chi^{\S}$ | Recalibrated<br>GND $\chi^{2\S\vert \vert }$ | ICI<sup>#</sup> | Recalibrated ICI<sup>#\|\|</sup> | Calibration<br>slope<sup>**</sup> |
| --- | --- | --- | --- | --- | --- | --- | --- |
| *C3PO* |  |  |  |  |  |  |  |
| PCE<br>(White women)* | 2.51 (2.43-2.59) | 0.768<br>(0.760-0.775) | 487, p<0.01 | 1689, p<0.01 | 0.018 (0.017-0.020)<br>p<0.01 | 0.034 (0.031-0.037)<br>p<0.01 | 0.67 (0.65-0.70)<br>p=0.02 |
| PCE<br>(Black women)* | 2.39 (2.17-2.64) | 0.724<br>(0.702-0.746) | 69, p<0.01 | 257, p<0.01 | 0.030 (0.023-0.036)<br>p<0.01 | 0.057 (0.050-0.064)<br>p<0.01 | 0.60 (0.53-0.67)<br>p=0.36 |
| PCE<br>(White men)* | 2.17 (2.11-2.24) | 0.738<br>(0.730-0.746) | 361, p<0.01 | 618, p<0.01 | 0.024 (0.022-0.027)<br>p<0.01 | 0.032 (0.029-0.035)<br>p<0.01 | 0.70 (0.68-0.73)<br>p=0.59 |
| PCE<br>(Black men)* | 2.04 (1.85-2.25) | 0.725<br>(0.698-0.751) | 21, p=0.02 | 18, p=0.03 | 0.012 (0-0.025)<br>p=0.06 | 0.010 (0-0.024)<br>p=0.83 | 0.88 (0.77-1.00)<br>p=0.84 |
| CHARGE-AF* | 2.56 (2.50-2.61) | 0.782<br>(0.777-0.787) | 1856, p<0.01 | 1367, p<0.01 | 0.028 (0.027-0.030)<br>p<0.01 | 0.019 (0.018-0.021)<br>p<0.01 | 0.77 (0.75-0.79)<br>p<0.01 |
| *Convenience Samples* |  |  |  |  |  |  |  |
| PCE<br>(White women)<sup>†</sup> | 2.44 (2.39-2.49) | 0.770<br>(0.764-0.775) | 1797, p<0.01 | 4923, p<0.01 | 0.032<br>(0.031-0.034) | 0.047<br>(0.044-0.049) | 0.64 (0.62-0.65) |
| PCE<br>(Black women)<sup>†</sup> | 2.29 (2.13-2.46) | 0.732<br>(0.716-0.748) | 213, p<0.01 | 562, p<0.01 | 0.046<br>(0.040-0.053) | 0.074<br>(0.067-0.081) | 0.56 (0.51-0.61) |
| PCE<br>(White men)<sup>†</sup> | 2.18 (2.13-2.22) | 0.744<br>(0.739-0.749) | 1291, p<0.01 | 1493, p<0.01 | 0.041<br>(0.038-0.043) | 0.039<br>(0.037-0.042) | 0.70 (0.68-0.71) |
| PCE<br>(Black men)<sup>†</sup> | 2.01 (1.88-2.15) | 0.727<br>(0.705-0.749) | 36, p<0.01 | 13, p=0.17 | 0.028<br>(0.018-0.037) | 0.012<br>(0.0026-0.022) | 0.87 (0.79-0.95) |
| CHARGE-AF<sup>†</sup> | 2.40 (2.38-2.43) | 0.781<br>(0.778-0.784) | 7188, p<0.01 | 8322, p<0.01 | 0.036<br>(0.035-0.036) | 0.028<br>(0.027-0.029) | 0.69 (0.68-0.70) |

\*PCE (White women): 4,231, 107,998, 7.1 (2.8, 10); PCE (Black women): 617, 8,450, 7.7 (2.9, 10); PCE (White men): 4,928, 76,304, 6.2 (2.3, 10); PCE (Black men): 425, 5,432, 6.7 (2.5, 10); CHARGE-AF: n events=7,877, N total=174,644, median follow-up, years (Q1,Q3): 5.0 (2.3,5.0)
†PCE (White women): 10,259, 182,349, 7.5 (3.6, 10); PCE (Black women): 1,119, 12,690, 7.2 (3.2, 10); PCE (White men): 12,891, 136,629, 6.2 (2.6, 10); PCE (Black men): 843, 8,558, 6.0 (2.6, 10); CHARGE-AF: n events=26,907, N total=501,272, median follow-up, years (Q1,Q3): 5.0 (2.0,5.0)
‡C-index calculated using the inverse probability of censoring weighting method<sup>24</sup>
§Greenwood-Nam-D'Agostino (GND) test, a test of calibration.<sup>26</sup> Lower chi-squared values suggest better calibration (across equally-sized samples). Significant p-values indicate evidence of miscalibration.
||Values after recalibration to the baseline hazard of the sample (see text)

We performed an analogous assessment of the PCE models within the MI/stroke Convenience Sample, which comprised 340,226 individuals with complete data to calculate the PCE. Compared to C3PO, individuals in the MI/stroke Convenience Sample had lower rates of cardiovascular comorbidity and anti-hypertensive medication use (**Table 1**). However, the observed 10-year MI/stroke risk was higher (cumulative risk of MI/stroke 10.6%, 95% CI 10.5-10.7; MI/stroke incidence rate 11.7 per 1,000 person-years, 95% CI 11.5-11.8). Cumulative risk curves demonstrated an abrupt rise in incident MI/stroke diagnoses shortly after the start of follow-up, which was not observed in C3PO (**Figure 5**). Discrimination of MI/stroke risk was similar to that observed in C3PO (c-index range 0.727-0.770, **Figure 6**), but calibration was worse (GND χ^2^ range 36-1,797; ICI range 0.028-0.046; calibration slope range 0.56-0.87, **Figure 7** and **Supplementary Figures 8-9**). Recalibration to the baseline hazard of the Convenience Sample did not correct miscalibration (GND χ^2^ range 13-4,923; ICI range 0.012-0.047, **Figure 7** and **Supplementary Figures 8-9**).

**Figure 5.**
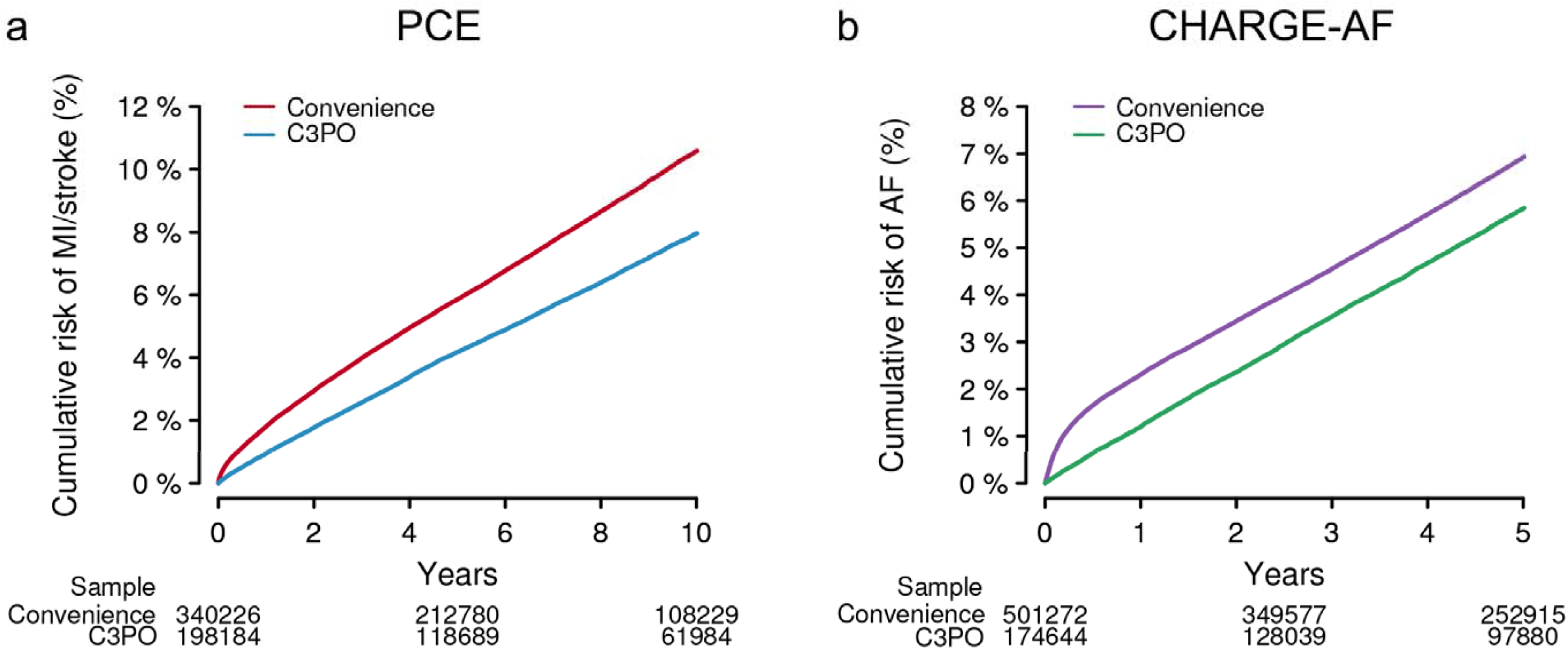
Cumulative event risk in C3PO vesus Convenience Samples. Depicted is Kaplan-Meier cumulative risk of MI/stroke (panel **a**) and AF (panel **b**) observed in C3PO (blue [left] and green [right]) versus the Convenience Samples (red [left] and purple [right). The number of individuals remaining at risk over time is labeled below each plot. Note an initial rapid inflection in MI/stroke and AF incidence observed in the Convenience Samples but not in C3PO.

**Figure 6.**
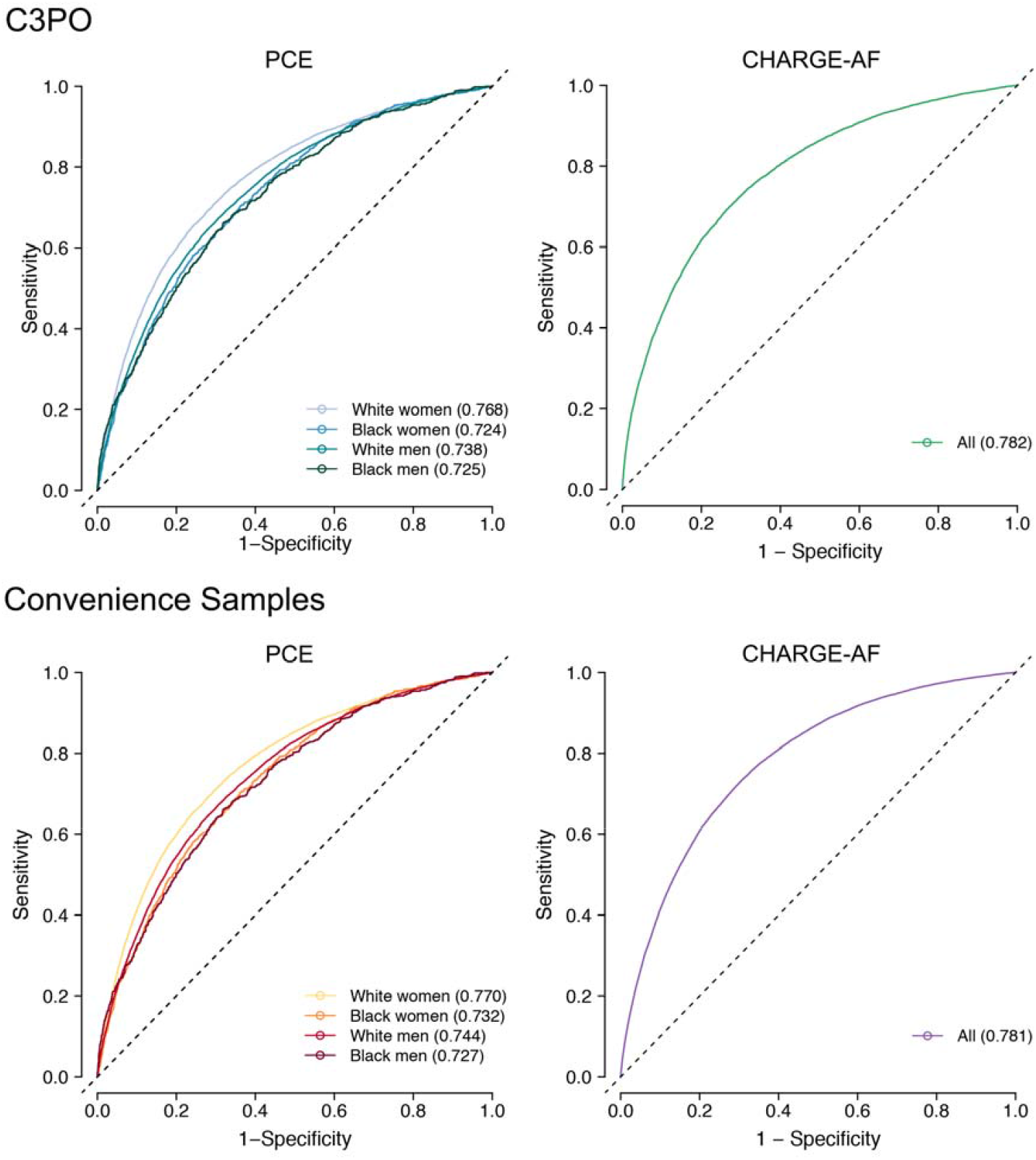
Model discrimination in C3PO and Convenience Samples. Depicted are time-dependent receiver operating characteristic curves for the Pooled Cohort Equations (PCE, left panels) and the CHARGE-AF score (right panels) in C3PO (top panels) versus the respective Convenience Samples (bottom panels). Each plot shows the discrimination performance of each risk score for its respective prediction target (i.e., 10-year MI/stroke for the PCE, 5-year incident AF for CHARGE-AF). Since the PCE score comprises four models stratified on the basis of sex and race, the curves for each score are represented separately (see legend). The c-index calculated using the inverse probability of censoring weighting method^24^ is depicted for each model.

**Figure 7.**
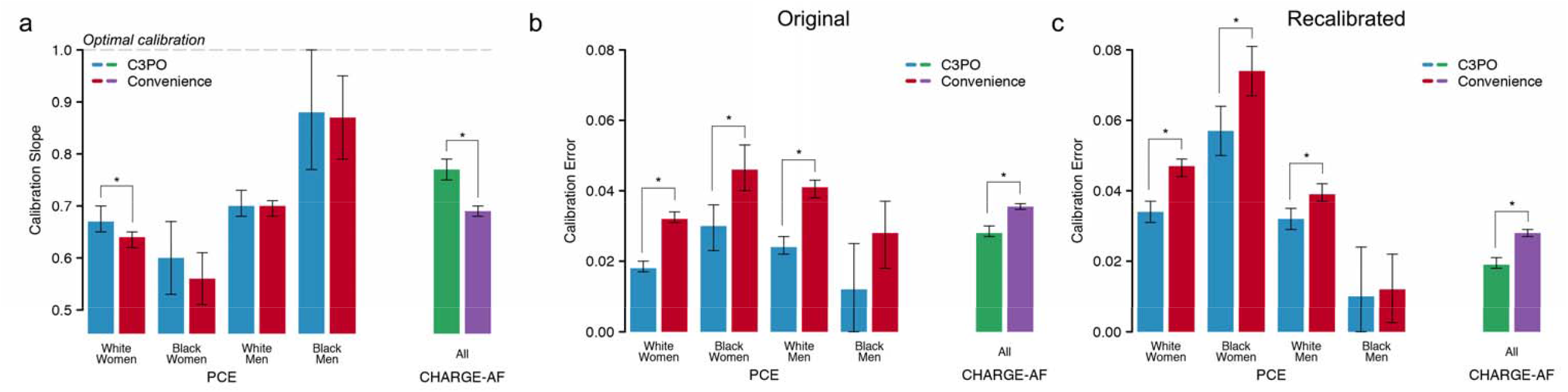
Model calibration in C3PO and Convenience Samples. Depicted is model calibration performance in C3PO versus the Convenience Samples. Panel **a** depicts the calibration slope for the PCE models (x-axis, left) and CHARGE-AF (x-axis, right) in C3PO (blue, green) versus the Convenience Samples (red, purple). The y-axis depicts the calibration slope, a measure of the relationship between predicted event risk and observed event incidence, where a slope of one indicates an optimal relationship (horizontal hashed line), with corresponding 95% confidence intervals. Panels **b** and **c** compare calibration error in C3PO versus the Convenience Samples. Calibration error is depicted on the y-axis using the Integrated Calibration Index (ICI, see text), where lower values indicate better absolute agreement between predicted risk and observed event incidence. Panel **b** depicts ICI values using the original models, while panel **c** depicts ICI values after recalibration to the baseline hazard of each sample. In all plots, statistically significant differences between values in C3PO versus the Convenience Sample (p<0.05) are depicted with an asterisk.

### AF analyses – CHARGE-AF

After excluding individuals for missing CHARGE-AF components, prevalent AF, absence of follow-up, or age outside 46-90 years, there were 174,644 individuals who were included in analyses of incident AF in C3PO (**Supplementary Figure 5**). Of the 174,644 individuals in the AF analysis, 38,528 (22.1%) would have been excluded in the absence of NLP-recovered data (**Figure 3**). At 5 years, there were 7,877 AF events (cumulative incidence 5.8%, 95% CI 5.7-6.0; AF incidence rate 12.1 per 1,000 person-years, 95% CI 11.8-12.3). The CHARGE-AF score was strongly associated with incident AF (hazard ratio [HR] per 1-standard deviation (SD) increase 2.56, 95% CI 2.50-2.61), with moderate discrimination (c-index 0.782, 95% 0.777-0.787), although CHARGE-AF substantially underestimated AF risk (GND χ^2^ 1,856, ICI 0.028, 95% CI 0.027-0.030). Calibration was much improved after recalibration to the baseline AF hazard in C3PO (GND χ^2^ 1,367; ICI 0.019, 95% CI 0.018-0.021; calibration slope 0.77, 95% CI 0.75-0.79). Details of CHARGE-AF model fit, discrimination, and calibration are shown in **Table 2**. The distribution of predicted AF risk before and after recalibration is shown in **Supplementary Figure 6**. Cumulative risk of AF stratified by predicted AF risk using the recalibrated CHARGE-AF score is shown in **Supplementary Figure 7**. Detailed assessments of CHARGE-AF calibration before and after recalibration are shown in **Supplementary Figures 8-9**. Model assessment with versus without NLP recovered values demonstrated similar performance but with greater precision in the NLP enabled analyses (**Supplementary Table 8**).

We performed an analogous assessment of CHARGE-AF within the AF Convenience Sample, which comprised 501,272 individuals with complete data to calculate the score. Similar to observations with MI/stroke, individuals in the AF Convenience Sample had lower rates of cardiovascular comorbidity and anti-hypertensive medication use (**Table 1**), yet higher 5-year AF risk (cumulative risk 6.9%, 95% CI 6.9-7.0; AF incidence rate 15.1 per 1,000 person-years, 95% CI 14.9-15.3). Cumulative risk curves again demonstrated an abrupt rise in incident AF diagnoses shortly after the start of follow-up, isolated to the Convenience Sample (**Figure 5**). Discrimination of AF risk using CHARGE-AF was similar to that observed in C3PO (c-index 0.781, 95% CI 0.778-0.784, **Figure 6**), but calibration was substantially worse (GND χ^2^ 7,188; ICI 0.036, 95% CI 0.035-0.036; calibration slope 0.69, 95% CI 0.68-0.70, p<0.01 for comparisons of ICI and calibration slope to C3PO, **Figure 7**). Calibration remained less favorable in the Convenience Sample even after recalibration to the baseline hazard (GND χ^2^ 8,322; ICI 0.028, 95% CI 0.027-0.029; **Figure 7** and **Supplementary Figures 8-9**).

## Discussion

In the present study, we demonstrate that selective sampling of individuals from a large multi-institutional EHR on the basis of longitudinal primary care encounters, and recovery of missingness using deep learning, enable EHR-based prediction with validity exceeding a conventional EHR based cohort sampling approach.^8,30,31^ C3PO comprises over a half-million individuals receiving longitudinal care over a decade of follow-up and, owing to the fact that it more closely mirrors the design of epidemiologic cohort studies as compared to conventional EHR based sampling, is likely to facilitate more generalizable insights. When compared to Convenience Samples derived from the same parent EHR with no requirement for longitudinal primary care, C3PO appeared less biased and offered greater data density. Leveraging neural network-based NLP models using unstructured notes, we achieved a roughly 30% reduction in missingness of baseline vital signs.

The JEDI data pipeline underlying C3PO, which we will make publicly available, provides a modular framework for processing and updating diverse EHR data in a manner conducive to an array of modeling approaches. We submit that JEDI, along with the principles underlying the development of C3PO, may enable future discovery by facilitating novel statistical and machine learning-based prediction and classification models utilizing diverse EHR data types available at scale and in a manner that reduces bias (**Figure 8**). The principles guiding the development of C3PO and the coding infrastructure for our analyses are widely extendable to external EHR data sets.

**Figure 8.**
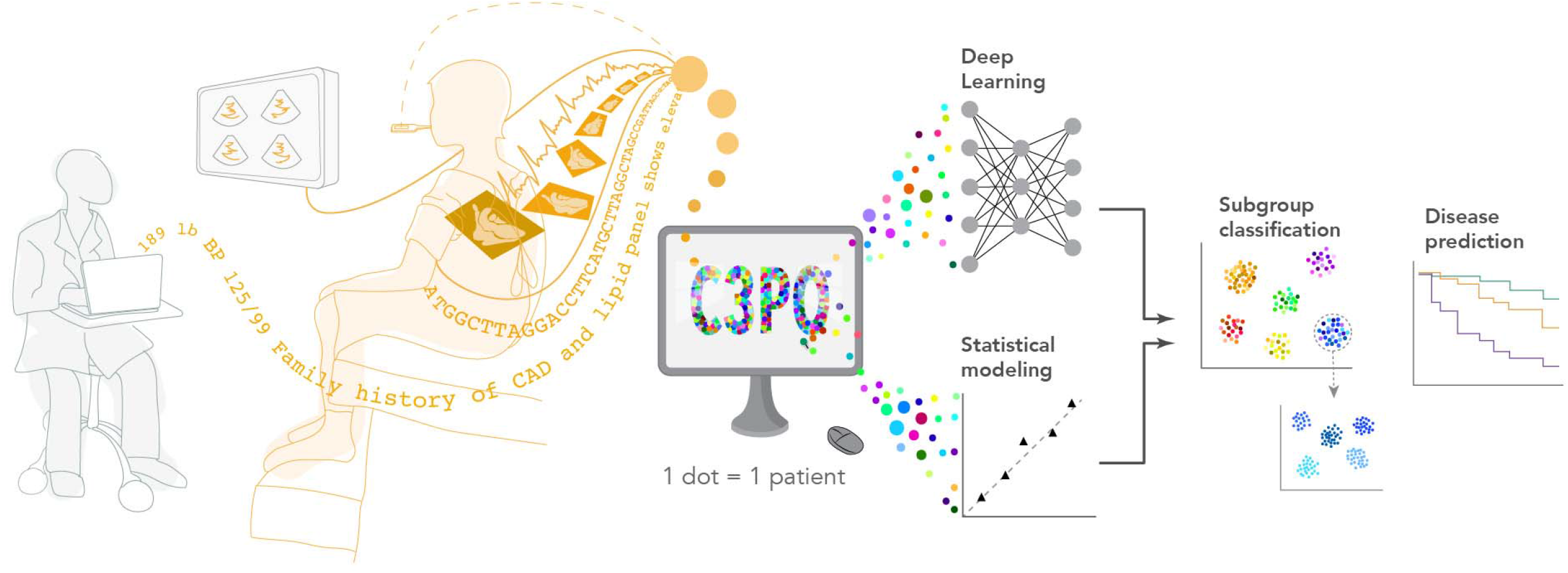
Conceptual overview of C3PO analysis methods. Depicted is a graphical overview of the potential analyses enabled by the Community Care Cohort Project (C3PO). By integrating diverse data types (e.g., diagnoses, imaging, vital signs, diagnostic test data, genetics), C3PO may enable methods such as traditional statistical modeling and deep learning to facilitate more accurate disease risk prediction models and enable deep phenotyping including disease subgroup identification.

The development of C3PO advances previous work leveraging EHR datasets to derive disease insights. The large scale and diversity of available features typically offered by EHR repositories is particularly conducive to the development of machine learning models. Recently, Artzi et al.^30^ developed a machine learning model trained on over a half-million individuals to predict the development of gestational diabetes. Chauhan et al.^32^ utilized an EHR dataset to develop a random forest-based prediction model to predict incident renal failure. EHR datasets have also been used to develop traditional statistical models, including risk scores for AF.^4,8^ In contrast to previous approaches, C3PO was specifically sampled in a manner intended to reduce selection bias inherent within most EHR-related study designs.

To that end, our observations suggest that EHR samples enriched for individuals receiving longitudinal primary care may offer a particularly efficient and valid setting for developing novel disease-related models. In the current study, we performed incident disease modeling using C3PO versus Convenience Samples including all individuals with complete data but with no requirement for longitudinal primary care. Both MI/stroke and AF incidence rates were higher in the Convenience Samples as opposed to C3PO, despite paradoxically lower rates of documented cardiovascular comorbidity. We suspect that selecting for individuals receiving longitudinal primary care improves detection of relevant clinical factors.

Similarly, we observed abrupt increases in incident diagnoses shortly after the start of follow-up in the Convenience Samples. By defining the start of follow-up as the second of the two qualifying PCP visits required for inclusion in C3PO, we submit there is greater likelihood for prevalent conditions to be appropriately recorded within the EHR prior to the onset of time-to-event analyses, minimizing misclassification of prevalent disease as compared to the Convenience Sample. Taken together, overall risk model performance was more consistent with expectations when deployed within C3PO as opposed to the Convenience Samples. Specifically, discrimination performance of the PCE and CHARGE-AF scores in C3PO was comparable to metrics reported in each score’s original validation study.^15,16^ Additionally, when compared to the Convenience Samples, model calibration was substantially favorable in C3PO, demonstrating a relationship between known risk factors and outcomes more consistent with prior evidence.^15,16^

We acknowledge that selecting a primary care population may introduce alternative biases (e.g., more likely to receive primary prevention or have insurance), which requires further study. Of note, EHR sample construction predicated on the needs of a specific analysis may also produce datasets that are less adaptable to other analytic frameworks.^10–12^ In contrast, the C3PO sampling design is readily amenable to an array of epidemiologic analyses (e.g., cross-sectional, retrospective cohort, case-control).

Our findings also imply that deep learning models applied on unstructured data have the potential to substantially reduce missingness in traditional EHR analyses. We leveraged neural network-based NLP methods, fine-tuned using relatively small amounts of labeled data,^33^ to accurately extract vital signs for an additional 80,000 individuals using unstructured text, reducing missingness by roughly one-third and increasing the precision of risk model performance estimates. We suspect that use of neural network-based approaches facilitated high accuracy despite a wide range of documentation patterns contained within notes spanning over 20 years – a substantial challenge to pattern matching approaches such as regular expressions. Importantly, when compared within individuals having features available from both tabular and NLP model sources, vital signs were consistently very highly correlated, with good agreement. We anticipate that analogous NLP models will be able to extract additional clinical parameters, such as laboratory values, which continue to exhibit considerable missingness in C3PO. Although we submit that recovery of actual data where possible is preferable to other methods of accounting for missingness, future work is needed to better understand the effects of incorporating features recovered from deep learning models on downstream analyses, and how such approaches differ from substitution methods such as multiple imputation.^34^

We submit that large and comprehensive EHR samples like C3PO have the potential to facilitate broad-ranging discovery leveraging diverse data types, provided that sufficient infrastructure exists to efficiently process, store, and analyze data within a unified framework. To that end, we have developed the JEDI pipeline, which automates processing and unification of diverse EHR data types within a harmonized, indexed file system amenable to a variety of statistical and machine learning-based approaches. Specifically, C3PO includes over 2.95 million ECGs, 450,000 echocardiograms, and millions of free text notes. Through linkage to the MGB Biobank biorepository, we anticipate that biological samples will be available within over 40,000 individuals. Facilitated by the JEDI pipeline, we expect that future models built within C3PO leveraging some or all of these data types will result in more accurate and generalizable disease prediction and classification models. Furthermore, recent work has suggested substantial value in the ability to utilize longitudinal EHR data to model patient trajectories.^35,36^ By providing nearly a decade of follow-up and a median of over 30 longitudinal office visits per person, we anticipate that C3PO will provide a very rich setting for trajectory modeling. Importantly, although the EHR data comprising C3PO is not sharable owing to concerns about data identifiability, the principles governing C3PO are widely applicable to EHR datasets and the JEDI pipeline will be publicly available to catalyze future research efforts related to the development of clinical models using rich and diverse EHR data.

Our study should be interpreted in the context of design. First, despite our purposeful intent to reduce bias by selecting individuals receiving regular primary care within our hospital network, residual indication bias is inevitable using EHR data. Nevertheless, we demonstrate that the approach taken to developing C3PO appears to reduce bias. Second, although we successfully employed NLP to reduce missingness rates for vital signs by roughly one-third, missingness of other features (e.g., cholesterol) remains considerable. We anticipate that similar NLP approaches will have utility in reducing missing data further. Third, although we utilized previously validated algorithms to define the presence of disease, some degree of misclassification of exposures and outcomes remains likely. Fourth, we identified individuals for inclusion in C3PO using EHR-based codes to identify office visits and a manually curated list of in-network primary care practice locations. Although two forms of validation support the accuracy of our selection methods, we acknowledge that the process is imperfect and would not easily extend to other EHRs. Fifth, most individuals included in C3PO are White, and therefore generalizability to populations with varying racial composition may be limited. However, we note that the absolute number of individuals of color within C3PO compares favorably to several other cohorts and EHR-based studies.^37–39^ Sixth, current results are observational and should not be used to infer causality.

In conclusion, we have developed C3PO, an EHR-based resource comprising over a half-million individuals within a large networked healthcare system. By sampling the full range of EHR data for individuals receiving regular primary care, EHR samples such as C3PO offer the potential to substantially reduce biases related to patient selection and missing data. By providing a broad array of data types, longitudinal measurements, and a flexible data structure conducive to multiple modeling frameworks, we anticipate that C3PO – and similarly constructed EHR datasets – will facilitate impactful discovery research.

## Disclosures

Dr. Philippakis receives sponsored research support from Bayer AG, IBM, Intel, and Verily. He has also received consulted fees from Novartis and Rakuten. He is a Venture Partner at GV and is compensated for this work. Dr. Ho receives sponsored research support from Bayer AG and Gilead Sciences. Dr. Ho has received research supplies from EcoNugenics. Dr. Friedman receives sponsored research support from Bayer AG and IBM. Dr. Anderson receives sponsored research support from Bayer AG and has consulted for ApoPharma and Invitae. Dr. Batra receives sponsored research support from Bayer AG and IBM, and consults for Novartis. Dr. Lubitz receives sponsored research support from Bristol Myers Squibb / Pfizer, Bayer AG, Boehringer Ingelheim, and Fitbit, and has consulted for Bristol Myers Squibb / Pfizer and Bayer AG, and participates in a research collaboration with IBM. Dr. Ellinor receives sponsored research support from Bayer AG and IBM Research and he has consulted for Bayer AG, Novartis, MyoKardia and Quest Diagnostics. Dr. Atlas receives sponsored research support from Bristol Myers Squibb / Pfizer and has consulted for Bristol Myers Squibb/Pfizer and Fitbit. Dr. Ashburner has received sponsored research support from Bristol Myers Squibb / Pfizer. Dr. Diedrich, Dr. Mielke, Dr. Eilken, Dr. Derix, and Ms. Ghadessi are employees of Bayer AG.

## Funding support

Dr. Khurshid is supported by NIH T32HL007208. Dr. Haimovich is supported by NIH R38HL150212. Dr. Atlas is supported by American Heart Association (AHA) grant 18SFRN34250007. Dr. Ashburner is supported by NIH K01HL148506 and AHA 18SFRN34250007. Dr. Ho is supported by NIH R01HL134893, R01HL140224, and K24HL153669. Dr. Lubitz is supported by NIH 1R01HL139731 and AHA 18SFRN34250007. Dr. Ellinor is supported by NIH 1R01HL092577, R01HL128914, K24HL105780, AHA 18SFRN34110082, and by the Foundation Leducq 14CVD01. Dr. Anderson is supported by NIH R01NS103924, U01NS069673, AHA 18SFRN34250007, and AHA-Bugher 21SFRN812095. This work was sponsored by Bayer AG.

## Notes

### Author Declarations

Study protocols complied with the tenets of the Declaration of Helsinki and were approved by the Mass General Brigham Institutional Review Board.

