## Supplemental Material for "Cohort Design and Natural Language Processing to Reduce Bias in Electronic Health Records Research: The Community Care Cohort Project"

### Supplementary Material

#### Index

**Methods 1**. Primary care algorithm validation

We then assessed the validity of our cohort construction process using two methods, namely manual chart review by two blinded clinicians (Validation 1), and assessment of overlap with an existing Massachusetts General Hospital (MGH)-based primary care registry (Validation 2). The validation was performed prior to requesting detailed data and prior to protected health information minimization (and restriction of the upper limit of age to <90 years), resulting in total n=523,445 for the candidate C3PO sample.

*Validation 1: Manual chart review*

Two clinical adjudicators blinded to C3PO selection algorithm status performed a manual chart review of 200 randomly selected algorithm-positive individuals ("algorithm-positive") and 200 randomly selected individuals with ≥1 office visit but none in a primary care location ("algorithm-negative"). A total of 60 algorithm-positive and 60 algorithm-negative records were overlapping between the adjudicated sets in order to assess inter-rater reliability.

In certain locations within MGB, the clinically accessible EHR lagged behind the availability of data sources from the data mart and RPDR. As a result, the ability to adjudicate the presence of primary care office visits in the earliest years of the C3PO cohort was limited. Frequently, there was indirect evidence of longitudinal primary care (e.g., notes making explicit mention of previous visits not available in the clinical EHR). As a result, we specified *a priori* two levels of adjudication for algorithm-positive individuals. In Tier 1, algorithm-positive individuals would be adjudicated as positive only if there was a narrative note confirming a primary care office visit (within ±7 days) on each of the two visit dates of interest. In Tier 2, algorithm-positive individuals would be adjudicated as positive if there was at least one pair of narrative notes confirming two primary care office visits 1-3 years apart (i.e., the inclusion criteria for C3PO) at any time in the individual's EHR history. In all cases, algorithm-negative individuals were adjudicated as correct if there was no evidence of a primary care office visit on both dates of interest.

The results of the adjudication process are summarized in **Supplemental Table 1**. Inter-rater agreement was excellent (kappa range 0.78-1). Both case and non-case algorithms met pre-specified criteria for sufficient algorithm accuracy (PPV ≥85%) to proceed with C3PO construction.

*Validation 2: Primary care network*

We assessed the validity of our cohort selection process by assessing overlap between the candidate C3PO cohort and an existing MGH-based primary care registry, to which we applied analogous selection methods. Specifically, we analyzed individuals who were represented in the MGH registry in ≥2 consecutive years between 2005-2017. Of 280,815 individuals in the MGH registry meeting the specified temporal selection criteria, the substantial majority (n=206,868; 73.7%) were represented in the candidate C3PO cohort (**Supplemental Figure 1**). The remaining discrepancy was attributed to differences in application of temporal selection criteria (1-3 year windows based on exact dates in C3PO, versus only calendar year data available in MGH registry), as well as exclusion of individuals aged <18 years at the start of follow-up in C3PO. Without application of temporal or age selection criteria, 277,780 out of 297,718 (93.3%) of the MGH registry was represented in C3PO (**Supplemental Figure 1**).

**Methods 2.** Construction of MI/stroke and AF Convenience Samples

To compare C3PO to EHR-based cohorts constructed using a more traditional analysis approach (i.e., selecting individuals solely on the basis of the availability of required data, with no need for longitudinal primary care), we developed MI/stroke and AF Convenience Samples derived from the same parent EHR data mart. From the source mart (N=3.6 million, see **Figure 1** in main manuscript), all individuals with available data for each component of the PCE (MI/stroke Convenience Sample) and each component of the CHARGE-AF score (AF Convenience Sample) were identified, with no requirement for primary care office visits. To maximize available sample size, we defined the start of follow-up for each individual as the earliest time at which all score components were available for that individual. We then excluded individuals who did not have all score components available within three years prior to each individuals' start follow-up date. We also excluded individuals with no follow-up data of any kind, as well as those having the relevant outcome at the start of follow-up. A summary of Convenience Sample construction is shown in **Supplemental Figure 3**. Disease-related exposure and outcome ascertainment was performed on the basis of at least one ICD, CPT, and/or EHR-specific diagnosis code present in the EHR mart corresponding to the relevant disease. Otherwise, all statistical analyses were analogous to the analyses described in the main text.

**Methods 3.** Natural language processing-based vital sign extraction

NLP methods have recently shown promising results for extracting information from unstructured text-based data.[1] Advanced neural network models such as Bidirectional Encoder Representations from Transformers (BERT)[2] have immense expressive power as their representations are derived from training on large corpora of text. These models can also be further pre-trained on domain-specific language, such as biomedical and clinical text. This has shown to improve performance on a number of clinical NLP tasks when compared to general language embeddings, including tasks like named entity recognition and diagnostic inference.[3,4] This bio-clinical-specific pretraining allows such models to be fine-tuned using relatively small amounts of "weakly" labeled data generated by a rule-based approach and perform exceptionally well on downstream tasks such as feature extraction from free note texts. In the current study, we utilized Bio+Discharge Summary BERT, a deep contextual word embedding model that has been pretrained consecutively on a large corpus of general English text (e.g., Wikipedia), biomedical text (PubMed abstracts and PubMed Central full-text articles),[4] and physician-written Discharge Summaries (from the MIMIC-III v1.4 database).[3,5]

We created a rule-based approach to automatically label the position of vital sign values in several different types of clinical notes. **Supplemental Table 3** demonstrates the context words, unit tokens and text patterns we considered for each feature, in addition to an example of labeled text in each case. For the purposes of training, four different note types were considered (e.g., discharge summaries, outpatient progress notes, inpatient progress notes) and 119,130 tokens were labeled as height, weight and blood pressure measurements and 61,646 tokens as corresponding units of height and weight. An additional 6,379 instances of vital signs were collected to be used as an independent test set. Training and test examples included 80-word spans surrounding the labeled tokens in order for the model to learn the context in which vital signs tend to appear. Bio+Discharge Summary BERT was fine-tuned for 5 epochs with the task of labeling the values identified by the rule-based approach.

We performed inference using this model on 9,522,262 notes for the 401,826 patients who had ≥1 eligible note within the three years prior to start of follow-up. We then performed the following post-processing on NLP extracted values:

1. Checked model identifications and extended identified tokens to include additional significant figures or unit tokens.
2. Harmonized model identifications into a single unit for each vital sign (i.e., kg for weight, cm for height, and mmHg for blood pressure) using a rule-based system to convert text patterns to numeric values.
3. Imposed physiological constraints for each feature, to ensure that each extraction was biologically plausible. The following constraints were used:
    a. 91 – 305 cm for height
    b. 20 – 450 kg for weight
    c. 50 – 300 mmHg for systolic, and 20 – 200 mmHg for diastolic blood pressure
4. Filtered out “optimal weights” that appeared in notes in addition to patient weights (a common model failure mode) using a regular expression that discarded weight identifications that were followed by variations of the phrase “for BMI of 25”.

**Supplemental Table 4** indicates NLP model yield and recovery rate for each vital sign. A study cardiologist then performed manual review of 50 randomly selected values for each vital sign (along with surrounding context from the note) obtained from an independent holdout set. All 200 values reviewed accurately represented the true vital sign of interest.

**Table 1**. Current procedural terminology codes corresponding to office visits

| **Code*** | **Description** |
|---|---|
| C99058 | Office services provided on an emergency basis |
| C99201 | New patient visit, level 1 |
| C99202 | New patient visit, level 2 |
| C99203 | New patient visit, level 3 |
| C99204 | New patient visit, level 4 |
| C99205 | New patient visit, level 5 |
| C99211 | Established patient visit, level 1 |
| C99212 | Established patient visit, level 2 |
| C99213 | Established patient visit, level 3 |
| C99214 | Established patient visit, level 4 |
| C99215 | Established patient visit, level 5 |
| CG0344 | Initial preventive physical examination |
| CG0438 | Annual wellness visit; initial |
| CG0439 | Annual wellness visit; subsequent |
| CG0463 | Hospital outpatient clinic visit |
| *Analogous institution-specific billing codes also included in the office visit definition | |

**Table 2.** Primary care algorithm validation results

| Sample | PPV (95% CI) | Raw Agreement | Kappa (95% CI) |
|---|---|---|---|
| Cases Tier 1 (n=200)* | 69.5 (62.6-75.8) | 59/60 (98.3%) | 0.96 (0.9-1) |
| Cases Tier 2 (n=200) † | 94.0 (89.8-96.7) | 58/60 (96.7%) | 0.78 (0.49-1) |
| Controls (n=200) | 91.5 (86.7-95.0) | 60/60 (100%) | 1 (1-1) |

*Defined as presence of directly verifiable primary care office visit on both qualifying dates (±7 days)
†Defined as presence of ≥2 directly verifiable primary care office visits within 1-3 consecutive years (C3PO temporal criteria), with allowance for misclassification of date

**Table 3**. Clinical factor definitions

| Phenotype | Code type | Data codes | Data code definitions |
|---|---|---|---|
| Diabetes | ICD9 | 249, 249.01, 249.1, 249.11, 249.2, 249.21, 249.3, 249.31, 249.4, 249.41, 249.5, 249.51, 249.6, 249.61, 249.7, 249.71, 249.8, 249.81, 249.9, 249.91, 250, 250.01, 250.02, 250.03, 250.1, 250.11, 250.12, 250.13, 250.2, 250.21, 250.22, 250.23, 250.3, 250.31, 250.32, 250.33, 250.4, 250.41, 250.42, 250.43, 250.5, 250.51, 250.52, 250.53, 250.6, 250.61, 250.62, 250.63, 250.7, 250.71, 250.72, 250.73, 250.8, 250.81, 250.82, 250.83, 250.9, 250.91, 250.92, 250.93, 357.2, 362.01, 362.02, 362.03, 362.04, 362.05, 362.06, 362.07, 366.41, 791.6 | Secondary diabetes mellitus without mention of complication, not stated as uncontrolled, or unspecified, Secondary diabetes mellitus without mention of complication, uncontrolled, Secondary diabetes mellitus with ketoacidosis, not stated as uncontrolled, or unspecified, Secondary diabetes mellitus with ketoacidosis, uncontrolled, Secondary diabetes mellitus with hyperosmolarity, not stated as uncontrolled, or unspecified, Secondary diabetes mellitus with hyperosmolarity, uncontrolled, Secondary diabetes mellitus with other coma, not stated as uncontrolled, or unspecified, Secondary diabetes mellitus with other coma, uncontrolled, Secondary diabetes mellitus with renal manifestations, not stated as uncontrolled, or unspecified, Secondary diabetes mellitus with renal manifestations, uncontrolled, Secondary diabetes mellitus with ophthalmic manifestations, not stated as uncontrolled, or unspecified, Secondary diabetes mellitus with ophthalmic manifestations, uncontrolled, Secondary diabetes mellitus with neurological manifestations, not stated as uncontrolled, or unspecified, Secondary diabetes mellitus with neurological manifestations, uncontrolled, Secondary diabetes mellitus with peripheral circulatory disorders, not stated as uncontrolled, or unspecified, Secondary diabetes mellitus with peripheral circulatory disorders, uncontrolled, Secondary diabetes mellitus with other specified manifestations, not stated as uncontrolled, or unspecified, Secondary diabetes mellitus with other specified manifestations, uncontrolled, Secondary diabetes mellitus with unspecified complication, not stated as uncontrolled, or unspecified, Secondary diabetes mellitus with unspecified complication, uncontrolled, Diabetes mellitus without mention of complication, type II or unspecified type, not stated as uncontrolled, Diabetes mellitus without mention of complication, type I [juvenile type], not stated as uncontrolled, Diabetes mellitus without mention of complication, type II or unspecified type, uncontrolled, Diabetes mellitus without mention of complication, type I [juvenile type], uncontrolled, Diabetes with ketoacidosis, type II or unspecified type, not stated as uncontrolled, Diabetes with ketoacidosis, type I [juvenile type], not stated as uncontrolled, Diabetes with ketoacidosis, type II or unspecified type, uncontrolled, Diabetes with ketoacidosis, type I [juvenile type], uncontrolled, Diabetes with hyperosmolarity, type II or unspecified type, not stated as uncontrolled, Diabetes with hyperosmolarity, type I [juvenile type], not stated as uncontrolled, Diabetes with hyperosmolarity, type II or unspecified type, uncontrolled, Diabetes with hyperosmolarity, type I [juvenile type], uncontrolled, Diabetes with other coma, type II or unspecified type, not stated as uncontrolled, Diabetes with other coma, type I [juvenile type], not stated as uncontrolled, Diabetes with other coma, type II or unspecified type, uncontrolled, Diabetes with other coma, type I [juvenile type], uncontrolled, Diabetes with renal manifestations, type II or unspecified type, not stated as uncontrolled, Diabetes with renal manifestations, type I [juvenile type], not stated as uncontrolled, Diabetes with renal manifestations, type II or unspecified type, uncontrolled, Diabetes with renal manifestations, type I [juvenile type], uncontrolled, Diabetes with ophthalmic manifestations, type II or unspecified type, not stated as uncontrolled, Diabetes with ophthalmic manifestations, type I [juvenile type], not stated as uncontrolled, Diabetes with ophthalmic manifestations, type II or unspecified type, uncontrolled, Diabetes with ophthalmic manifestations, type I [juvenile type], uncontrolled, Diabetes with |

| Phenotype | Code type | Data codes | Data code definitions |
|---|---|---|---|
| | | | neurological manifestations, type II or unspecified type, not stated as uncontrolled, Diabetes with neurological manifestations, type I [juvenile type], not stated as uncontrolled, Diabetes with neurological manifestations, type II or unspecified type, uncontrolled, Diabetes with neurological manifestations, type I [juvenile type], uncontrolled, Diabetes with peripheral circulatory disorders, type II or unspecified type, not stated as uncontrolled, Diabetes with peripheral circulatory disorders, type I [juvenile type], not stated as uncontrolled, Diabetes with peripheral circulatory disorders, type II or unspecified type, uncontrolled, Diabetes with peripheral circulatory disorders, type I [juvenile type], uncontrolled, Diabetes with other specified manifestations, type II or unspecified type, not stated as uncontrolled, Diabetes with other specified manifestations, type I [juvenile type], not stated as uncontrolled, Diabetes with other specified manifestations, type II or unspecified type, uncontrolled, Diabetes with other specified manifestations, type I [juvenile type], uncontrolled, Diabetes with unspecified complication, type II or unspecified type, not stated as uncontrolled, Diabetes with unspecified complication, type I [juvenile type], not stated as uncontrolled, Diabetes with unspecified complication, type II or unspecified type, uncontrolled, Diabetes with unspecified complication, type I [juvenile type], uncontrolled, Polyneuropathy in diabetes, Background diabetic retinopathy, Proliferative diabetic retinopathy, Nonproliferative diabetic retinopathy NOS, Mild nonproliferative diabetic retinopathy, Moderate nonproliferative diabetic retinopathy, Severe nonproliferative diabetic retinopathy, Diabetic macular edema, Diabetic cataract, Acetonuria |
| Diabetes | ICD10 | E08.00, E08.01, E08.10, E08.11, E08.21, E08.22, E08.29, E08.311, E08.319, E08.321, E08.329, E08.331, E08.339, E08.341, E08.349, E08.351, E08.359, E08.36, E08.39, E08.40, E08.41, E08.42, E08.43, E08.44, E08.49, E08.51, E08.52, E08.610, E08.618, E08.620, E08.621, E08.622, E08.628, E08.630, E08.638, E08.641, E08.649, E08.65, E08.69, E08.8, E08.9, E08.90, E09.00, E09.01, E09.10, E09.11, E09.21, E09.22, E09.29, E09.311, E09.319, E09.321, E09.329, | Diabetes Mellitus Due To Underlying Condition With Hyperosmolarity without nonketotic hyperglycemic-hyperosmolar coma (NKHHC), Diabetes Mellitus Due To Underlying Condition With Hyperosmolarity With Coma, Diabetes Mellitus Due To Underlying Condition With Ketoacidosis Without Coma, Diabetes Mellitus Due To Underlying Condition With Ketoacidosis With Coma, Diabetes Mellitus Due To Underlying Condition With Diabetic Nephropathy, Diabetes Mellitus Due To Underlying Condition With Diabetic chronic kidney disease, Diabetes Mellitus Due to underlying condition with other diabetic kidney complication, Diabetes Mellitus Due To Underlying Condition With Unspecified Diabetic Retinopathy With Macular Edema, Diabetes Mellitus Due To Underlying Condition With Unspecified Diabetic Retinopathy Without Macular Edema, Diabetes Mellitus due to underlying condition with mild nonproliferative diabetic retinopathy with macular edema, Diabetes mellitus due to underlying condition with mild nonproliferative diabetic retinopathy without macular edema, Diabetes mellitus due to underlying condition with moderate nonproliferative diabetic retinopathy with macular edema, Diabetes mellitus due to underlying condition with moderate nonproliferative diabetic retinopathy without macular edema, Diabetes mellitus due to underlying condition with severe nonproliferative diabetic retinopathy with macular edema, Diabetes mellitus due to underlying condition with severe nonproliferative diabetic retinopathy without macular edema, Diabetes mellitus due to underlying condition with proliferative diabetic retinopathy with macular edema, Diabetes mellitus due to underlying condition with proliferative diabetic retinopathy without macular edema, Diabetes Mellitus Due To Underlying Condition With Diabetic Cataract, Diabetes Mellitus Due To Underlying Condition With Other Diabetic Ophthalmic Complication, Diabetes Mellitus Due To Underlying Condition With Diabetic Neuropathy, Unspecified, Diabetes Mellitus Due To Underlying Condition With Diabetic Mononeuropathy, Diabetes Mellitus Due To Underlying Condition With Diabetic Polyneuropathy, Diabetes Mellitus Due To Underlying Condition With Diabetic Autonomic (Poly)Neuropathy, Diabetes Mellitus Due To Underlying |

| Phenotype | Code type | Data codes | Data code definitions |
|---|---|---|---|
| | | E09.331, E09.339, E09.341, E09.349, E09.351, E09.359, E09.36, E09.39, E09.40, E09.41, E09.42, E09.43, E09.44, E09.49, E09.51, E09.52, E09.59, E09.610, E09.618, E09.620, E09.621, E09.622, E09.628, E09.630, E09.638, E09.641, E09.649, E09.65, E09.69, E09.8, E09.9, E10.10, E10.11, E10.21, E10.22, E10.29, E10.311, E10.319, E10.321, E10.329, E10.331, E10.339, E10.341, E10.349, E10.351, E10.359, E10.36, E10.39, E10.40, E10.41, E10.42, E10.43, E10.44, E10.49, E10.51, E10.52, E10.59, E10.610, E10.618, E10.620, E10.621, E10.622, E10.628, E10.630, E10.638, E10.641, E10.649, E10.65, E10.69, E10.8, E10.9, E11.00, E11.01, E11.21, E11.22, E11.29, E11.311, E11.319, E11.321, E11.329, E11.331, E11.339, E11.341, E11.349, E11.351, E11.359, E11.36, E11.39, E11.40, E11.41, E11.42, E11.51, | Condition With Diabetic Amyotrophy, Diabetes Mellitus Due To Underlying Condition With Other Diabetic Neurological Complication, Diabetes Mellitus Due To Underlying Condition With Diabetic Peripheral Angiopathy Without Gangrene, Diabetes Mellitus Due To Underlying Condition With Diabetic Neuropathic Arthropathy, Diabetes Mellitus Due To Underlying Condition with diabetic neuropathic arthropathy, Diabetes Mellitus Due To Underlying Condition With Diabetic arthropathy, Diabetes Mellitus Due To Underlying Condition With diabetic dermatitis, Diabetes Mellitus Due To Underlying Condition With Foot Ulcer, Diabetes Mellitus Due To Underlying Condition With Other Skin ulcer, Diabetes Mellitus Due To Underlying Condition With other skin complications, Diabetes Mellitus Due To Underlying Condition With periodontal disease, Diabetes Mellitus Due To Underlying Condition With other oral complications, Diabetes Mellitus Due To Underlying Condition With hypoglycemia with coma, Diabetes mellitus due to underlying condition with hypoglycemia without coma, Diabetes Mellitus Due To Underlying Condition With Hyperglycemia, Diabetes Mellitus Due To Underlying Condition With Other Specified Complication, Diabetes Mellitus Due To Underlying Condition With Unspecified Complications, Diabetes mellitus due to underlying condition without Complications, Diabetes Mellitus Due To Underlying Condition Without Complications, Drug or Chemical induced diabetes mellitus with hyperosmolarity without nonketotic hyperglycemic-hyperosmolar coma (NKHHC), Drug Or Chemical Induced Diabetes Mellitus With Hyperosmolarity With Coma, Drug Or Chemical Induced Diabetes Mellitus With Ketoacidosis Without Coma, Drug Or Chemical Induced Diabetes Mellitus With Ketoacidosis With Coma, Drug Or Chemical Induced Diabetes Mellitus With Diabetic Nephropathy, Drug Or Chemical Induced Diabetes Mellitus With diabetic chronic kidney disease, Drug or chemical induced diabetes mellitus with other diabetic kidney complication, Drug Or Chemical Induced Diabetes Mellitus With Unspecified Diabetic Retinopathy With Macular Edema, Drug Or Chemical Induced Diabetes Mellitus With Unspecified Diabetic Retinopathy Without Macular Edema, Drug or chemical induced diabetes mellitus with mild nonproliferative diabetic retinopathy with macular edema, Drug or chemical induced diabetes mellitus with mild nonproliferative diabetic retinopathy without macular edema, Drug or chemical induced diabetes mellitus with moderate nonproliferative diabetic retinopathy with macular edema, Drug or chemical induced diabetes mellitus with moderate nonproliferative diabetic retinopathy without macular edema, Drug or chemical induced diabetes mellitus with severe nonproliferative diabetic retinopathy with macular edema, Drug or chemical induced diabetes mellitus with severe nonproliferative diabetic retinopathy without macular edema, Drug or chemical induced diabetes mellitus with proliferative diabetic retinopathy with macular edema, Drug or chemical induced diabetes mellitus with proliferative diabetic retinopathy without macular edema, Drug Or Chemical Induced Diabetes Mellitus With Diabetic Cataract, Drug Or Chemical Induced Diabetes Mellitus With Other Diabetic Ophthalmic Complication, Drug Or Chemical Induced Diabetes Mellitus With Neurological Complications With Diabetic Neuropathy, Unspecified, Drug Or Chemical Induced Diabetes Mellitus With Neurological Complications With Diabetic Mononeuropathy, Drug Or Chemical Induced Diabetes Mellitus With Neurological Complications With Diabetic Polyneuropathy, Drug Or Chemical Induced Diabetes Mellitus With Neurological Complications With Diabetic Autonomic (Poly)Neuropathy, Drug Or Chemical Induced Diabetes Mellitus With Neurological Complications With Diabetic Amyotrophy, Drug Or Chemical Induced Diabetes Mellitus With Neurological Complications With Other Diabetic Neurological Complication, Drug Or Chemical Induced Diabetes Mellitus |

| Phenotype | Code type | Data codes | Data code definitions |
|---|---|---|---|
| | | E11.52, E11.59, E11.610, E11.618, E11.620, E11.621, E11.622, E11.628, E11.630, E11.638, E11.641, E11.649, E11.65, E11.69, E11.8, E11.9, E13.00, E13.01, E13.10, E13.11, E13.21, E13.22, E13.29, E13.311, E13.319, E13.321, E13.329, E13.331, E13.339, E13.341, E13.349, E13.351, E13.359, E13.36, E13.39, E13.40, E13.41, E13.42, E13.43, E13.44, E13.49, E13.51, E13.52, E13.59, E13.610, E13.618, E13.620, E13.621, E13.622, E13.628, E13.630, E13.638, E13.641, E13.649, E13.65, E13.69, E13.8, E13.9, R82.4 | With Diabetic Peripheral Angiopathy Without Gangrene, Drug Or Chemical Induced Diabetes Mellitus With Diabetic peripheral angiopathy with gangrene, Drug or chemical induced diabetes mellitus with other circulatory complications, Drug Or Chemical Induced Diabetes Mellitus With Diabetic neuropathic Arthropathy, Drug or chemical induced diabetes mellitus with other diabetic arthropathy, Drug Or Chemical Induced Diabetes Mellitus With diabetic dermatitis, Drug Or Chemical Induced Diabetes Mellitus With foot ulcer, Drug Or Chemical Induced Diabetes Mellitus With Other Skin ulcer, Drug Or Chemical Induced Diabetes Mellitus With other skin complications, Drug Or Chemical Induced Diabetes Mellitus With periodontal disease, Drug Or Chemical Induced Diabetes Mellitus With other oral complications, Drug Or Chemical Induced Diabetes Mellitus With Hypoglycemia with coma, Drug Or Chemical Induced Diabetes Mellitus With hypoglycemia without coma, Drug Or Chemical Induced Diabetes Mellitus With Hyperglycemia, Drug Or Chemical Induced Diabetes Mellitus With Other Specified Complication, Drug Or Chemical Induced Diabetes Mellitus With Unspecified Complications, Drug Or Chemical Induced Diabetes Mellitus without complications, Type 1 Diabetes Mellitus With Ketoacidosis Without Coma, Type 1 Diabetes Mellitus With Ketoacidosis With Coma, Type 1 Diabetes Mellitus With Diabetic Nephropathy, Type 1 Diabetes Mellitus With diabetic chronic kidney disease, Type 1 Diabetes Mellitus With other diabetic kidney complication, Type 1 Diabetes Mellitus With Unspecified Diabetic Retinopathy With Macular Edema, Type 1 Diabetes Mellitus With Unspecified Diabetic Retinopathy Without Macular Edema, Type 1 Diabetes mellitus with mild nonproliferative diabetic retinopathy with macular edema, Type 1 Diabetes mellitus with mild nonproliferative diabetic retinopathy without macular edema, Type 1 diabetes mellitus with moderate nonproliferative diabetic retinopathy with macular edema, Type 1 diabetes mellitus with moderate nonproliferative diabetic retinopathy without macular edema, Type 1 diabetes mellitus with severe nonproliferative diabetic retinopathy with macular edema, Type 1 diabetes mellitus with severe nonproliferative diabetic retinopathy without macular edema, Type 1 diabetes mellitus with proliferative diabetic retinopathy with macular edema, Type 1 diabetes mellitus with proliferative diabetic retinopathy without macular edema, Type 1 Diabetes Mellitus With Diabetic Cataract, Type 1 Diabetes Mellitus With Other Diabetic Ophthalmic Complication, Type 1 Diabetes Mellitus With Diabetic Neuropathy, Unspecified, Type 1 Diabetes Mellitus With diabetic mononeuropathy, Type 1 Diabetes Mellitus With Diabetic polyneuropathy, Type 1 Diabetes mellitus with diabetic autonomic (poly)neuropathy, Type 1 diabetes mellitus with diabetic amyotrophy, Type 1 diabetes mellitus with other diabetic neurological complication, Type 1 Diabetes Mellitus With Diabetic Peripheral Angiopathy Without Gangrene, Type 1 diabetes mellitus with diabetic peripheral angiopathy with gangrene, Type 1 diabetes mellitus with other circulatory complications, Type 1 diabetes mellitus with diabetic neuropathic arthropathy, Type 1 Diabetes Mellitus With Diabetic arthropathy, Type 1 Diabetes Mellitus With diabetic dermatitis, Type 1 Diabetes Mellitus With Other foot ulcer, Type 1 Diabetes Mellitus With Other Skin ulcer, Type 1 Diabetes Mellitus With other skin complications, Type 1 Diabetes Mellitus With periodontal disease, Type 1 Diabetes Mellitus With other oral complications, Type 1 Diabetes Mellitus With Hypoglycemia With Coma, Type 1 Diabetes Mellitus With Hypoglycemia without coma, Type 1 Diabetes Mellitus With Hyperglycemia, Type 1 Diabetes Mellitus With Other Specified Complication, Type 1 Diabetes Mellitus With Unspecified Complications, Type 1 Diabetes Mellitus Without Complications, Type 2 Diabetes Mellitus With Hyperosmolarity Without Nonketotic |

| Phenotype | Code type | Data codes | Data code definitions |
|---|---|---|---|
| | | | Hyperglycemic-Hyperosmolar Coma (Nkhhc), Type 2 Diabetes Mellitus With Hyperosmolarity With Coma, Type 2 diabetes mellitus with diabetic nephropathy, Type 2 diabetes mellitus with diabetic chronic kidney disease, Type 2 diabetes mellitus with other diabetic kidney complication, Type 2 Diabetes Mellitus With Unspecified Diabetic Retinopathy With Macular Edema, Type 2 Diabetes Mellitus With Unspecified Diabetic Retinopathy Without Macular Edema, Type 2 Diabetes Mellitus With Mild Nonproliferative Diabetic Retinopathy With Macular Edema, Type 2 Diabetes Mellitus With Mild Nonproliferative Diabetic Retinopathy Without Macular Edema, Type 2 Diabetes Mellitus With Moderate Nonproliferative Diabetic Retinopathy With Macular Edema, Type 2 Diabetes Mellitus with moderate nonproliferative diabetic retinopathy Without Macular Edema, Type 2 Diabetes Mellitus With Severe Nonproliferative Diabetic Retinopathy With Macular Edema, Type 2 Diabetes Mellitus With severe nonproliferative Diabetic Retinopathy Without Macular Edema, Type 2 Diabetes Mellitus with proliferative diabetic retinopathy with macular edema, Type 2 Diabetes Mellitus With proliferative diabetic retinopathy without macular edema, Type 2 Diabetes Mellitus With Diabetic Cataract, Type 2 Diabetes Mellitus With Other Diabetic Ophthalmic Complication, Type 2 Diabetes Mellitus With Diabetic Neuropathy, Unspecified, Type 2 Diabetes Mellitus With Diabetic mononeuropathy, Type 2 Diabetes Mellitus with diabetic polyneuropathy, Type 2 Diabetes Mellitus With Diabetic Peripheral Angiopathy Without Gangrene, Type 2 Diabetes Mellitus With diabetic peripheral Angiopathy With Gangrene, Type 2 Diabetes Mellitus with other circulatory complications, Type 2 diabetes mellitus with diabetic neuropathic arthropathy, Type 2 diabetes mellitus with other diabetic arthropathy, Type 2 Diabetes Mellitus with diabetic dermatitis, Type 2 Diabetes Mellitus With foot ulcer, Type 2 Diabetes Mellitus With Other Skin ulcer, Type 2 Diabetes Mellitus With other skin complications, Type 2 Diabetes Mellitus With periodontal disease, Type 2 Diabetes Mellitus With other oral complications, Type 2 Diabetes Mellitus With Hypoglycemia With Coma, Type 2 Diabetes Mellitus With Hypoglycemia without coma, Type 2 Diabetes Mellitus With Hyperglycemia, Type 2 Diabetes Mellitus With Other Specified Complication, Type 2 Diabetes Mellitus With Unspecified Complications, Type 2 Diabetes Mellitus Without Complications, Other specified diabetes mellitus with hyperosmolarity without nonketotic hyperglycemic-hyperosmolar coma (NKHHC), Other specified diabetes mellitus with hyperosmolarity with coma, Other specified diabetes mellitus with ketoacidosis without coma, Other specified diabetes mellitus with ketoacidosis with coma, Other specified diabetes mellitus with diabetic nephropathy, Other specified diabetes mellitus with diabetic chronic kidney disease, Other specified diabetes mellitus with other diabetic kidney complication, Other specified diabetes mellitus with unspecified diabetic retinopathy with macular edema, Other specified diabetes mellitus with unspecified diabetic retinopathy without macular edema, Other specified diabetes mellitus with mild nonproliferative diabetic retinopathy with macular edema, Other specified diabetes mellitus with mild nonproliferative diabetic retinopathy without macular edema, Other specified diabetes mellitus with moderate nonproliferative diabetic retinopathy with macular edema, Other specified diabetes mellitus with moderate nonproliferative diabetic retinopathy without macular edema, Other specified diabetes mellitus with severe nonproliferative diabetic retinopathy with macular edema, Other specified diabetes mellitus with severe nonproliferative diabetic retinopathy without macular edema, Other specified diabetes mellitus with proliferative diabetic retinopathy with macular edema, Other specified diabetes mellitus with proliferative diabetic retinopathy without macular edema, Other Specified Diabetes Mellitus With diabetic cataract, Other |

| Phenotype | Code type | Data codes | Data code definitions |
|---|---|---|---|
| | | | Specified Diabetes Mellitus With other diabetic ophthalmic complication, Other Specified Diabetes Mellitus With Diabetic neuropathy, unspecified, Other Specified Diabetes Mellitus With Diabetic mononeuropathy, Other Specified Diabetes Mellitus With Diabetic Polyneuropathy, Other Specified Diabetes Mellitus With Diabetic Autonomic (Poly)Neuropathy, Other Specified Diabetes Mellitus With Diabetic Amyotrophy, Other Specified Diabetes Mellitus With Other Diabetic Neurological Complication, Other specified diabetes mellitus with diabetic peripheral angiopathy without gangrene, Other specified diabetes mellitus with diabetic peripheral angiopathy with gangrene, Other Specified Diabetes Mellitus With Other Circulatory Complications, Other specified diabetes mellitus with diabetic neuropathic arthropathy, Other specified diabetes mellitus with other diabetic arthropathy, Other Specified Diabetes Mellitus With Diabetic Dermatitis, Other Specified Diabetes Mellitus With Foot Ulcer, Other Specified Diabetes Mellitus With Other Skin Ulcer, Other specified diabetes mellitus with other skin complications, Other specified diabetes mellitus with periodontal disease, Other specified diabetes mellitus with other oral complications, Other Specified Diabetes Mellitus With Hypoglycemia With Coma, Other Specified Diabetes Mellitus With Hypoglycemia Without Coma, Other Specified Diabetes Mellitus With Hyperglycemia, Other Specified Diabetes Mellitus With Other Specified Complication, Other Specified Diabetes Mellitus With Unspecified Complications, Other Specified Diabetes Mellitus Without Complications, Acetonuria |
| Diabetes | Med | metformin, glucophage, riomet, fortamet, glumetzia, chlorpropamide, glimepiride, glyburide, glipizide, tolazamide, tolbutamide, diabinese, amaryl, diabeta, micronase, glucotrol, glynase, tolinase, orinase, tolbutamide, repaglinide, nateglinide, prandin, starlix, pioglitazone, rosiglitazone, actos, avandia, sitagliptin, saxagliptin, linagliptin, alogliptin, januvia, onglyza, tradjenta, nesina, acarbose, miglitol, precose, glyset, pramlintide, symlin, liraglutide, exenatide, | Biguanides, Sulfonylureas, Meglitinides, Thiazolidinediones, DPP-4 Inhibitors, Alpha-glucosidase inhibitors, Amylin analogue, GLP-1 Agonist, SGLT2 Inhibitor, Combination pills, long-actin insulin |

| Phenotype | Code type | Data codes | Data code definitions |
|---|---|---|---|
| | | albiglutide, dulaglutide, victoza, bydureon, byetta, tanzeum, trulicity, canagliflozin, dapagliflozin, empagliflozin, invokana, farxiga, jardiance, actoplus, glucovance, metaglip, janumet, kombiglyze, prandimet, duetact, kazano, invokamet, xigduo, synjardy, jentadueto, avandamet, oseni, glyxambi, avandaryl, juvisync, glargine, basaglar, lantus, toujeo, detemir, degludec, levemir, tresiba | |
| Hyperlipidemia | ICD9 | 272, 272.1, 272.2, 272.3, 272.4, 272.5, 272.6, 272.7, 272.8, 272.9, 759.9 | Pure hypercholesterolemia, Pure hyperglyceridemia, Mixed hyperlipidemia, Hyperchylomicronemia, Other hyperlipidemia, Hyperlipidemia, unspecified, Lipoprotein deficiency, Lipidoses, Other specified metabolic disorders, Disorder of lipoprotein metabolism, unspecified, Disorders of bile acid and cholesterol metabolism |
| Hyperlipidemia | ICD10 | E71.30, E75.21, E75.22, E75.5, E75.6, E77.0, E77.1, E78.0, E78.1, E78.2, E78.3, E78.4, E78.5, E78.6, E78.7, E78.70, E78.79, E78.81, E78.89, E78.9, E88.1, E88.89 | Disorder of fatty-acid metabolism, unspecified, Lipidoses, Lipidoses, Other lipid storage disorders, Lipid storage disorder, unspecified, Lipidoses, Lipidoses, Pure hypercholesterolemia, Pure hyperglyceridemia, Mixed hyperlipidemia, Hyperchylomicronemia, Other hyperlipidemia, Hyperlipidemia, unspecified, Lipoprotein deficiency, Disorders of bile acid and cholesterol metabolism, Disorder of bile acid and cholesterol metabolism, unspecified, Other disorders of bile acid and cholesterol metabolism, Lipid dermatoarthritis, Other lipoprotein metabolism disorders, Disorder of lipoprotein metabolism, unspecified, Lipodystrophy, Other specified metabolic disorders |
| Hyperlipidemia | Med | advicor, alirocumab, altoprev, antara, atorvastatin, cholestyramine, colesevelam, colestid, colestipol, crestor, evolocumab, ezetimibe, fenofibrate, fenofibric | |

| Phenotype | Code type | Data codes | Data code definitions |
|---|---|---|---|
| | | acid, fluvastatin, gemfibrozil, juxtapid, kynamro, lescol, lipitor, livalo, lomitapide, lopid, lovastatin, mipomersen, niacin, niacor, niaspan, nicotinic acid, pitavastatin, praluent, pravastatin, prevachol, prevalite, repatha, rosuvastatin, simvastatin, tricor, triglide, trilipix, vytorin, welchol, zetia, zocor | |
| Heart Failure | ICD9 | 398.91, 402.01, 402.11, 402.91, 404.01, 404.03, 404.11, 404.13, 404.91, 404.93, 428, 428.1, 428.2, 428.21, 428.22, 428.23, 428.3, 428.31, 428.32, 428.33, 428.4, 428.41, 428.42, 428.43, 428.9 | Rheumatic heart failure (congestive), Malignant hypertensive heart disease with heart failure, Benign hypertensive heart disease with heart failure, Unspecified hypertensive heart disease with heart failure , Hypertensive heart and chronic kidney disease, malignant, with heart failure and with chronic kidney disease stage I through stage IV, or unspecified, Hypertensive heart and chronic kidney disease, malignant, with heart failure and with chronic kidney disease stage V or end stage renal disease, Hypertensive heart and chronic kidney disease, benign, with heart failure and with chronic kidney disease stage I through stage IV, or unspecified, Hypertensive heart and chronic kidney disease, benign, with heart failure and chronic kidney disease stage V or end stage renal disease, Hypertensive heart and chronic kidney disease, unspecified, with heart failure and with chronic kidney disease stage I through stage IV, or unspecified, Hypertensive heart and chronic kidney disease, unspecified, with heart failure and chronic kidney disease stage V or end stage renal disease, Congestive heart failure, unspecified, Left heart failure, Systolic heart failure, unspecified, Acute systolic heart failure, Chronic systolic heart failure, Acute on chronic systolic heart failure, Diastolic heart failure, unspecified, Acute diastolic heart failure, Chronic diastolic heart failure, Acute on chronic diastolic heart failure, Combined systolic and diastolic heart failure, unspecified, Acute combined systolic and diastolic heart failure, Chronic combined systolic and diastolic heart failure, Acute on chronic combined systolic and diastolic heart failure, Heart failure, unspecified |
| Heart Failure | ICD10 | I09.81, I11.0, I13.0, I13.2, I50.1, I50.20, I50.21, I50.22, I50.23, I50.30, I50.31, I50.32, I50.33, I50.40, I50.41, I50.42, I50.43, I50.9, I97.130, I97.131 | Rheumatic Heart Failure, Hypertensive Heart Disease With Heart Failure, Hypertensive Heart And Chronic Kidney Disease With Heart Failure And Stage 1 Through Stage 4 Chronic Kidney Disease, Or Unspecified Chronic Kidney Disease, Hypertensive Heart And Chronic Kidney Disease With Heart Failure And With Stage 5 Chronic Kidney Disease, Or End Stage Renal Disease, Left Ventricular Failure, Unspecified Systolic (Congestive) Heart Failure, Acute Systolic (Congestive) Heart Failure, Chronic Systolic (Congestive) Heart Failure, Acute On Chronic Systolic (Congestive) Heart Failure, Unspecified Diastolic (Congestive) Heart Failure, Acute Diastolic (Congestive) Heart Failure, Chronic Diastolic (Congestive) Heart Failure, Acute On Chronic Diastolic (Congestive) Heart Failure, Unspecified Combined Systolic (Congestive) And Diastolic (Congestive) Heart Failure, Acute Combined Systolic (Congestive) And Diastolic (Congestive) |

| Phenotype | Code type | Data codes | Data code definitions |
|---|---|---|---|
| | | | Heart Failure, Chronic Combined Systolic (Congestive) And Diastolic (Congestive) Heart Failure, Acute On Chronic Combined Systolic (Congestive) And Diastolic (Congestive) Heart Failure, Heart Failure, Unspecified, Postprocedural heart failure following cardiac surgery, Postprocedural heart failure following other surgery |
| Coronary Heart Disease | ICD9 | 411, 411.1, 411.81, 411.89, 413, 413.9, 414, 414.01, 414.02, 414.03, 414.04, 414.05, 414.06, 414.07, 414.1, 414.11, 414.19, 414.2, 414.3, 414.4, 414.8, 414.9, 429.2, 996.03, V45.81, V45.82 | Postmyocardial infarction syndrome, Intermediate coronary syndrome, Acute coronary occlusion without myocardial infarction, Other acute and subacute forms of ischemic heart disease, other, Angina decubitus, Other and unspecified angina pectoris, Coronary atherosclerosis of unspecified type of vessel, native or graft, Coronary atherosclerosis of native coronary artery, Coronary atherosclerosis of autologous vein bypass graft, Coronary atherosclerosis of nonautologous biological bypass graft, Coronary atherosclerosis of artery bypass graft, Coronary atherosclerosis of unspecified bypass graft, Coronary atherosclerosis of native coronary artery of transplanted heart, Coronary atherosclerosis of bypass graft (artery) (vein) of transplanted heart, Aneurysm of heart (wall), Aneurysm of coronary vessels, Chronic ischemic heart disease, unspecified, Chronic total occlusion of coronary artery, Coronary atherosclerosis due to lipid rich plaque, Coronary atherosclerosis due to calcified coronary lesion, Other specified forms of chronic ischemic heart disease, Chronic ischemic heart disease, unspecified, Cardiovascular disease, unspecified, Mechanical complication due to coronary bypass graft, Aortocoronary bypass status, Percutaneous transluminal coronary angioplasty status |
| Coronary Heart Disease | ICD10 | I20.0, I20.1, I20.8, I20.9, I21.01, I21.02, I21.09, I21.11, I21.19, I21.21, I21.29, I21.3, I21.4, I22.0, I22.1, I22.2, I22.8, I22.9, I23.0, I23.1, I23.2, I23.3, I23.4, I23.5, I23.6, I23.7, I23.8, I24.0, I24.8, I25.10, I25.110, I25.111, I25.118, I25.119, I25.3, I25.41, I25.42, I25.6, I25.700, I25.701, I25.708, I25.709, I25.710, I25.711, I25.718, I25.719, I25.720, I25.721, I25.728, I25.729, I25.730, I25.731, I25.738, I25.739, I25.790, I25.791, I25.798, I25.799, I25.810, I25.811, | Unstable angina, Angina pectoris with documented spasm, Other forms of angina pectoris, Angina pectoris, unspecified, ST elevation (STEMI) myocardial infarction involving left main coronary artery, ST elevation (STEMI) myocardial infarction involving left anterior descending coronary artery, ST elevation (STEMI) myocardial infarction involving other coronary artery of anterior wall, ST elevation (STEMI) myocardial infarction involving right coronary artery, ST elevation (STEMI) myocardial infarction involving other coronary artery of inferior wall, ST elevation (STEMI) myocardial infarction involving left circumflex coronary artery, ST elevation (STEMI) myocardial infarction involving other sites, ST elevation (STEMI) myocardial infarction of unspecified site , Non-ST elevation (NSTEMI) myocardial infarction , Subsequent ST elevation (STEMI) myocardial infarction of anterior wall, Subsequent ST elevation (STEMI) myocardial infarction of inferior wall, Subsequent non-ST elevation (NSTEMI) myocardial infarction , Subsequent ST elevation (STEMI) myocardial infarction of other sites, Subsequent ST elevation (STEMI) myocardial infarction of unspecified site, Hemopericardium as current complication following acute myocardial infarction, Atrial septal defect as current complication following acute myocardial infarction, Ventricular septal defect as current complication following acute myocardial infarction, Rupture of cardiac wall without hemopericardium as current complication following acute myocardial infarction, Rupture of chordae tendineae as current complication following acute myocardial infarction, Rupture of papillary muscle as current complication following acute myocardial infarction, Thrombosis of atrium, auricular appendage, and ventricle as current complications following acute myocardial infarction, Postinfarction angina, Other current complications following acute myocardial infarction, Acute coronary thrombosis not resulting in myocardial infarction, Other forms of acute ischemic heart disease , Atherosclerotic heart disease of native coronary artery without angina pectoris, Atherosclerotic heart disease of native coronary artery with unstable angina pectoris, Atherosclerotic heart disease of native coronary artery with angina pectoris with documented spasm, |

| Phenotype | Code type | Data codes | Data code definitions |
|---|---|---|---|
| | | I25.812, I25.82, I25.83, I25.84, I25.89, I25.9, Z95.1, Z98.61 | Atherosclerotic heart disease of native coronary artery with other forms of angina pectoris, Atherosclerotic heart disease of native coronary artery with unspecified angina pectoris, Aneurysm of heart, Coronary artery aneurysm, Coronary artery dissection, Silent myocardial ischemia, Atherosclerosis of coronary artery bypass graft(s), unspecified, with unstable angina pectoris, Atherosclerosis of coronary artery bypass graft(s), unspecified, with angina pectoris with documented spasm, Atherosclerosis of coronary artery bypass graft(s), unspecified, with other forms of angina pectoris, Atherosclerosis of coronary artery bypass graft(s), unspecified, with unspecified angina pectoris, Atherosclerosis of autologous vein coronary artery bypass graft(s) with unstable angina pectoris, Atherosclerosis of autologous vein coronary artery bypass graft(s) with angina pectoris with documented spasm, Atherosclerosis of autologous vein coronary artery bypass graft(s) with other forms of angina pectoris, Atherosclerosis of autologous vein coronary artery bypass graft(s) with unspecified angina pectoris, Atherosclerosis of autologous artery coronary artery bypass graft(s) with unstable angina pectoris, Atherosclerosis of autologous artery coronary artery bypass graft(s) with angina pectoris with documented spasm, Atherosclerosis of autologous artery coronary artery bypass graft(s) with other forms of angina pectoris, Atherosclerosis of autologous artery coronary artery bypass graft(s) with unspecified angina pectoris, Atherosclerosis of nonautologous biological coronary artery bypass graft(s) with unstable angina pectoris, Atherosclerosis of nonautologous biological coronary artery bypass graft(s) with angina pectoris with documented spasm, Atherosclerosis of nonautologous biological coronary artery bypass graft(s) with other forms of angina pectoris, Atherosclerosis of nonautologous biological coronary artery bypass graft(s) with unspecified angina pectoris, Atherosclerosis of other coronary artery bypass graft(s) with unstable angina pectoris, Atherosclerosis of other coronary artery bypass graft(s) with angina pectoris with documented spasm, Atherosclerosis of other coronary artery bypass graft(s) with other forms of angina pectoris, Atherosclerosis of other coronary artery bypass graft(s) with unspecified angina pectoris, Atherosclerosis of coronary artery bypass graft(s) without angina pectoris , Atherosclerosis of native coronary artery of transplanted heart without angina pectoris, Atherosclerosis of bypass graft of coronary artery of transplanted heart without angina pectoris, Chronic total occlusion of coronary artery, Coronary atherosclerosis due to lipid rich plaque , Coronary atherosclerosis due to calcified coronary lesion, Other forms of chronic ischemic heart disease, Chronic ischemic heart disease, unspecified, Presence of aortocoronary bypass graft, Coronary angioplasty status |
| Hypertension | ICD9 | 401, 401.1, 401.9, 402, 402.01, 402.1, 402.11, 402.9, 402.91, 403, 403.01, 403.1, 403.11, 403.9, 403.91, 404, 404.01, 404.02, 404.03, 404.1, 404.11, 404.12, 404.13, 404.9, 404.91, 404.92, 404.93, 405.01, 405.09, 405.11, 405.19, | Malignant essential hypertension, Benign essential hypertension, Unspecified essential hypertension, Malignant hypertensive heart disease without heart failure, Malignant hypertensive heart disease with heart failure, Benign hypertensive heart disease without heart failure, Benign hypertensive heart disease with heart failure, Unspecified hypertensive heart disease without heart failure, Unspecified hypertensive heart disease with heart failure, Hypertensive chronic kidney disease, malignant, with chronic kidney disease stage I through stage IV, or unspecified, Hypertensive chronic kidney disease, malignant, with chronic kidney disease stage V or end stage renal disease, Hypertensive chronic kidney disease, benign, with chronic kidney disease stage I through stage IV, or unspecified, Hypertensive chronic kidney disease, benign, with chronic kidney disease stage V or end stage renal disease, Hypertensive chronic kidney disease, unspecified, with chronic kidney disease stage I through stage IV, or unspecified, Hypertensive chronic kidney disease, unspecified, with chronic kidney disease stage V or end stage renal disease, |

| Phenotype | Code type | Data codes | Data code definitions |
|---|---|---|---|
| | | 405.91, 405.99, 437.2, 796.2 | Hypertensive heart and chronic kidney disease, malignant, without heart failure and with chronic kidney disease stage I through stage IV, or unspecified, Hypertensive heart and chronic kidney disease, malignant, with heart failure and with chronic kidney disease stage I through stage IV, or unspecified, Hypertensive heart and chronic kidney disease, malignant, without heart failure and with chronic kidney disease stage V or end stage renal disease, Hypertensive heart and chronic kidney disease, malignant, with heart failure and with chronic kidney disease stage V or end stage renal disease, Hypertensive heart and chronic kidney disease, benign, without heart failure and with chronic kidney disease stage I through stage IV, or unspecified, Hypertensive heart and chronic kidney disease, benign, with heart failure and with chronic kidney disease stage I through stage IV, or unspecified, Hypertensive heart and chronic kidney disease, benign, without heart failure and with chronic kidney disease stage V or end stage renal disease, Hypertensive heart and chronic kidney disease, benign, with heart failure and chronic kidney disease stage V or end stage renal disease, Hypertensive heart and chronic kidney disease, unspecified, without heart failure and with chronic kidney disease stage I through stage IV, or unspecified, Hypertensive heart and chronic kidney disease, unspecified, with heart failure and with chronic kidney disease stage I through stage IV, or unspecified, Hypertensive heart and chronic kidney disease, unspecified, without heart failure and with chronic kidney disease stage V or end stage renal disease, Hypertensive heart and chronic kidney disease, unspecified, with heart failure and chronic kidney disease stage V or end stage renal disease, Malignant renovascular hypertension, Other malignant secondary hypertension, Benign renovascular hypertension, Other benign secondary hypertension, Unspecified renovascular hypertension, Other unspecified secondary hypertension, Hypertensive encephalopathy, Elevated blood pressure reading without diagnosis of hypertension |
| Hypertension | ICD10 | I10, I11.0, I11.9, I12.0, I12.9, I13.0, I13.10, I13.11, I13.2, I15.0, I15.1, I15.2, I15.8, I15.9 | Essential (Primary) Hypertension, Hypertensive Heart Disease with Heart Failure, Hypertensive Heart Disease without Heart Failure, Hypertensive Chronic Kidney Disease with Stage 5 Chronic Kidney Disease or end stage renal disease, Hypertensive Chronic Kidney Disease with Stage 1 through Stage 4 Chronic Kidney Disease, or unspecified chronic kidney disease, Hypertensive heart and chronic kidney disease with heart failure and stage 1 through stage 4 chronic kidney disease, or unspecified chronic kidney disease , Hypertensive heart and chronic kidney disease without heart failure, with stage 1 through stage 4 chronic kidney disease, or unspecified chronic kidney disease , Hypertensive heart and chronic kidney disease without heart failure, with stage 5 chronic kidney disease, or end stage renal disease, Hypertensive heart and chronic kidney disease with heart failure and with stage 5 chronic kidney disease, or end stage renal disease, Renovascular Hypertension, Hypertension secondary to other renal disorders, Hypertension secondary to endocrine disorders, Other Secondary Hypertension, Secondary hypertension, unspecified |
| Valvular Disease | ICD9 | 35.05, 35.06, 35.1, 35.11, 35.12, 35.13, 35.14, 35.2, 35.21, 35.22, 35.23, 35.24, 35.25, 35.26, 35.27, 35.28, 35.96, 394, 394.1, 394.2, 394.9, | Endovascular replacement of aortic valve, Transapical replacement of aortic valve, Open heart valvuloplasty without replacement, unspecified valve, Open heart valvuloplasty of aortic valve without replacement, Open heart valvuloplasty of mitral valve without replacement, Open heart valvuloplasty of pulmonary valve without replacement, Open heart valvuloplasty of tricuspid valve without replacement, Open and other replacement of unspecified heart valve, Open and other replacement of aortic valve with tissue graft, Open and other replacement of aortic valve, Open and other replacement of mitral valve with tissue graft, Open and other replacement of mitral valve, Open and other replacement of pulmonary valve with tissue graft, Open and |

| Phenotype | Code type | Data codes | Data code definitions |
|---|---|---|---|
| | | 396, 396.1, 396.2, 396.3, 396.8, 396.9, V42.2, V43.3 | other replacement of pulmonary valve, Open and other replacement of tricuspid valve with tissue graft, Open and other replacement of tricuspid valve, Percutaneous balloon valvuloplasty, Mitral stenosis, Rheumatic Mitral Insufficiency, Mitral stenosis with insufficiency, Other unspecified mitral valve disease, Mitral valve stenosis and aortic valve stenosis, Mitral valve stenosis and aortic valve insufficiency, Mitral valve insufficiency and aortic valve stenosis, Mitral valve insufficiency and aortic valve insufficiency, Multiple involvement of mitral and aortic valves, Mitral and aortic valve diseases, unspecified, Heart valve replaced by transplant, Heart valve replaced by other means |
| Valvular Disease | ICD10 | I05.0, I05.1, I05.2, I05.8, I05.9, I06.8 , I06.9, I07.8, I07.9, I08.0, I08.1, I08.3, I08.8, I08.9, I09.1, I34.0, I34.1, I34.2, I34.8, I34.9, I35.0, I35.1, I35.2, I35.8, I35.9, I36.0, I36.1, I36.2, I36.8, I36.9, I37.0, I37.1, I37.2, I37.8, I37.9, I38 | Rheumatic Mitral Stenosis, Rheumatic Mitral Insufficiency, Rheumatic Mitral Stenosis With Insufficiency, Other Rheumatic Mitral Valve Diseases, Rheumatic mitral valve disease, unspecified, Other rheumatic aortic valve diseases, Rheumatic aortic valve disease, unspecified, Other rheumatic tricuspid valve diseases, Rheumatic tricuspid valve disease, unspecified, Rheumatic Disorders Of Both Mitral And Aortic Valves, Rheumatic Disorders Of Both Mitral And tricuspid Valves, Combined rheumatic disorders of mitral, aortic, and tricuspid valves, Other Rheumatic Multiple Valve Diseases, Rheumatic Multiple Valve Disease, Unspecified, Rheumatic diseases of endocardium, valve unspecified, Nonrheumatic mitral (valve) insufficiency, Nonrheumatic mitral (valve) prolapse, Nonrheumatic mitral (valve) stenosis, Other nonrheumatic mitral valve disorders, Nonrheumatic mitral valve disorder, unspecified, Nonrheumatic aortic (valve) stenosis, Nonrheumatic aortic (valve) insufficiency, Nonrheumatic aortic (valve) stenosis with insufficiency, Other nonrheumatic aortic valve disorders, Nonrheumatic aortic valve disorder, unspecified, Nonrheumatic tricuspid (valve) stenosis, Nonrheumatic tricuspid (valve) insufficiency, Nonrheumatic tricuspid (valve) stenosis with insufficiency, Other nonrheumatic tricuspid valve disorders, Nonrheumatic tricuspid valve disorder, unspecified, Nonrheumatic pulmonary valve stenosis, Nonrheumatic pulmonary valve insufficiency, Nonrheumatic pulmonary valve stenosis with insufficiency, Other nonrheumatic pulmonary valve disorders, Nonrheumatic pulmonary valve disorder, unspecified, Endocarditis, valve unspecified |
| Valvular Disease | CPT | 33400, 33401, 33403, 33405, 33406, 33411, 33412, 33413, 33420, 33422, 33425, 33426, 33427, 33430, 33606, 33611, 33612, 33645, 33665, 33670, 33681, 33684, 33688, 33690 | Repair Of Aortic Valve, Valvuloplasty, Open, Valvuloplasty, W/Cp Bypass, Replacement Of Aortic Valve, Replacement Of Aortic Valve, Replacement Of Aortic Valve, Replacement Of Aortic Valve, Replacement Of Aortic Valve, Revision Of Mitral Valve, Revision Of Mitral Valve, Repair Of Mitral Valve, Repair Of Mitral Valve, Repair Of Mitral Valve, Replacement Of Mitral Valve, Anastomosis/artery-aorta, Repair double ventricle , Repair double ventricle , Revision of heart veins , Repair of heart defects, Repair of heart chambers , Repair heart septum defect , Repair heart septum defect , Repair heart septum defect , Reinforce pulmonary artery |
| Stroke/TIA | ICD9 | 362.31, 362.32, 362.33, 362.34, 388.02, 430, 431, 432.9, 433.01, 433.11, 433.21, 433.31, 433.81, 433.91, 434, 434.01, 434.1, 434.11, 434.9, 434.91, 435, | Central retinal artery occlusion, Retinal arterial branch occlusion, Partial retinal arterial occlusion, Transient retinal arterial occlusion, Transient ischemic deafness, Subarachnoid hemorrhage, Intracerebral hemorrhage, Unspecified intracranial hemorrhage, Occlusion and stenosis of basilar artery with cerebral infarction, Occlusion and stenosis of carotid artery with cerebral infarction, Occlusion and stenosis of vertebral artery with cerebral infarction, Occlusion and stenosis of multiple and bilateral precerebral arteries with cerebral infarction, Occlusion and stenosis of other specified precerebral artery with cerebral infarction, Occlusion and stenosis of unspecified precerebral artery with cerebral infarction, Cerebral thrombosis |

| Phenotype | Code type | Data codes | Data code definitions |
|---|---|---|---|
| | | 435.1, 435.2, 435.3, 435.8, 435.9, 437.1, 437.7, 437.9, 438.1, 438.11, 438.12, 438.13, 438.14, 438.2, 438.21, 438.22, 438.81, 438.82, 438.83, 438.89, 438.9, 997.02, V12.54 | without mention of cerebral infarction, Cerebral thrombosis with cerebral infarction, Cerebral embolism without mention of cerebral infarction, Cerebral embolism with cerebral infarction, Cerebral artery occlusion, unspecified without mention of cerebral infarction, Cerebral artery occlusion, unspecified with cerebral infarction, Basilar artery syndrome, Vertebral artery syndrome , Subclavian steal syndrome, Vertebrobasilar artery syndrome, Other specified transient cerebral ischemias, Unspecified transient cerebral ischemia, Other generalized ischemic cerebrovascular disease, Transient global amnesia, Unspecified cerebrovascular disease, Late effects of cerebrovascular disease, speech and language deficit, unspecified, Late effects of cerebrovascular disease, aphasia, Late effects of cerebrovascular disease, dysphasia, Late effects of cerebrovascular disease, dysarthria, Late effects of cerebrovascular disease, fluency disorder, Late effects of cerebrovascular disease, hemiplegia affecting unspecified side, Late effects of cerebrovascular disease, hemiplegia affecting dominant side, Late effects of cerebrovascular disease, hemiplegia affecting nondominant side, Other late effects of cerebrovascular disease, apraxia, Other late effects of cerebrovascular disease, dysphagia, Other late effects of cerebrovascular disease, facial weakness, Other late effects of cerebrovascular disease, Unspecified late effects of cerebrovascular disease, Iatrogenic cerebrovascular infarction or hemorrhage, Personal history of transient ischemic attack (TIA), and cerebral infarction without residual deficits |
| Stroke/TIA | IDC10 | G45.0, G45.1, G45.2, G45.3, G45.4, G45.8, G46.3, G46.4, H34.00, H34.01, H34.02, H34.03, H34.10, H34.11, H34.12, H34.13, H34.211, H34.212, H34.213, H34.219, H34.231, H34.232, H34.233, H34.239, H93.099, I60.9, I61.9, I62.9, I63.00, I63.011, I63.012, I63.019, I63.111, I63.112, I63.119, I63.12, I63.131, I63.132, I63.139, I63.19, I63.20, I63.211, I63.212, I63.219, I63.22, I63.231, I63.232, I63.239, I63.29, I63.30, I63.311, I63.312, I63.319, I63.321, I63.322, I63.329, | Vertebro-Basilar Artery Syndrome, Carotid Artery Syndrome, Multiple and bilateral precerebral artery syndromes, Amaurosis fugax, Transient Global Amnesia, Other transient cerebral ischemic attacks and related syndromes, Brain stem stroke syndrome, Cerebellar stroke syndrome, Transient Retinal Artery Occlusion, Unspecified Eye, Transient retinal artery occlusion, right eye, Transient retinal artery occlusion, left eye, Transient retinal artery occlusion, bilateral, Central retinal artery occlusion, unspecified eye, Central retinal artery occlusion, right eye, Central retinal artery occlusion, left eye, Central retinal artery occlusion, bilateral, Partial retinal artery occlusion, right eye, Partial retinal artery occlusion, left eye, Partial retinal artery occlusion, bilateral, Partial Retinal Artery Occlusion, Unspecified Eye, Retinal artery branch occlusion, right eye, Retinal artery branch occlusion, left eye, Retinal artery branch occlusion, bilateral, Retinal Artery Branch Occlusion, Unspecified Eye, Unspecified Degenerative and Vascular Disorders of Unspecified Ear, Nontraumatic Subarachnoid Hemorrhage, Unspecified, Nontraumatic intracerebral Hemorrhage, unspecified, Nontraumatic Intracranial Hemorrhage, Unspecified, Cerebral infarction due to thrombosis of unspecified precerebral artery, Cerebral infarction due to thrombosis of right vertebral artery, Cerebral infarction due to thrombosis of left vertebral artery, Cerebral infarction due to thrombosis of unspecified vertebral artery, Cerebral infarction due to embolism of right vertebral artery, Cerebral infarction due to embolism of left vertebral artery, Cerebral infarction due to embolism of unspecified vertebral artery , Cerebral infarction due to embolism of basilar artery, Cerebral infarction due to embolism of right carotid artery, Cerebral infarction due to embolism of left carotid artery, Cerebral infarction due to embolism of unspecified carotid artery, Cerebral infarction due to embolism of other precerebral artery, Cerebral infarction due to unspecified occlusion or stenosis of unspecified precerebral arteries , Cerebral infarction due to unspecified occlusion or stenosis of right vertebral arteries, Cerebral infarction due to unspecified occlusion or stenosis of left vertebral arteries, Cerebral infarction due to unspecified occlusion or stenosis of unspecified vertebral arteries, Cerebral Infraction Due to Unspecified Occlusion or Stenosis of Basilar |

| Phenotype | Code type | Data codes | Data code definitions |
|---|---|---|---|
| | | I63.331, I63.332, I63.339, I63.341, I63.342, I63.349, I63.40, I63.411, I63.412, I63.419, I63.421, I63.422, I63.429, I63.431, I63.432, I63.439, I63.49, I63.50, I63.511, I63.512, I63.519, I63.521, I63.522, I63.529, I63.531, I63.532, I63.539, I63.541, I63.542, I63.549, I63.59, I63.6, I63.8, I63.9, I66.01, I66.02, I66.03, I66.09, I66.11, I66.12, I66.13, I66.19, I66.21, I66.22, I66.23, I66.29, I66.3, I66.8, I66.9, I67.81, I67.82, I67.841, I67.848, I67.89, I67.9, I69.80, I69.81, I69.820, I69.821, I69.822, I69.823, I69.828, I69.831, I69.832, I69.833, I69.834, I69.839, I69.841, I69.842, I69.843, I69.844, I69.849, I69.851, I69.852, I69.853, I69.854, I69.859, I69.861, I69.862, I69.863, I69.864, I69.865, I69.869, I69.890, I69.891, I69.892, I69.893, I69.898, I69.90, I69.91, I69.920, I69.921, | Arteries, Cerebral infarction due to unspecified occlusion or stenosis of right carotid arteries, Cerebral infarction due to unspecified occlusion or stenosis of left carotid arteries, Cerebral infarction due to unspecified occlusion or stenosis of unspecified carotid arteries, Cerebral infarction due to unspecified occlusion or stenosis of other precerebral arteries, Cerebral infarction due to thrombosis of unspecified cerebral artery , Cerebral infarction due to thrombosis of right middle cerebral artery, Cerebral infarction due to thrombosis of left middle cerebral artery, Cerebral infarction due to thrombosis of unspecified middle cerebral artery, Cerebral infarction due to thrombosis of right anterior cerebral artery, Cerebral infarction due to thrombosis of left anterior cerebral artery, Cerebral infarction due to thrombosis of unspecified anterior cerebral artery, Cerebral infarction due to thrombosis of right posterior cerebral artery, Cerebral infarction due to thrombosis of left posterior cerebral artery, Cerebral infarction due to thrombosis of unspecified posterior cerebral artery, Cerebral infarction due to thrombosis of right cerebellar artery, Cerebral infarction due to thrombosis of left cerebellar artery, Cerebral infarction due to thrombosis of unspecified cerebellar artery, Cerebral infarction due to embolism of unspecified cerebral artery , Cerebral infarction due to embolism of right middle cerebral artery, Cerebral infarction due to embolism of left middle cerebral artery, Cerebral infarction due to embolism of unspecified middle cerebral artery, Cerebral infarction due to embolism of right anterior cerebral artery, Cerebral infarction due to embolism of left anterior cerebral artery, Cerebral infarction due to embolism of unspecified anterior cerebral artery, Cerebral infarction due to embolism of right posterior cerebral artery, Cerebral infarction due to embolism of left posterior cerebral artery, Cerebral infarction due to embolism of unspecified posterior cerebral artery , Cerebral infarction due to embolism of other cerebral artery, Cerebral infarction due to unspecified occlusion or stenosis of unspecified cerebral artery , Cerebral infarction due to unspecified occlusion or stenosis of right middle cerebral artery, Cerebral infarction due to unspecified occlusion or stenosis of left middle cerebral artery, Cerebral infarction due to unspecified occlusion or stenosis of unspecified middle cerebral artery, Cerebral infarction due to unspecified occlusion or stenosis of right anterior cerebral artery, Cerebral infarction due to unspecified occlusion or stenosis of left anterior cerebral artery, Cerebral infarction due to unspecified occlusion or stenosis of unspecified anterior cerebral artery, Cerebral infarction due to unspecified occlusion or stenosis of right posterior cerebral artery, Cerebral infarction due to unspecified occlusion or stenosis of left posterior cerebral artery, Cerebral infarction due to unspecified occlusion or stenosis of unspecified posterior cerebral artery, Cerebral infarction due to unspecified occlusion or stenosis of right cerebellar artery, Cerebral infarction due to unspecified occlusion or stenosis of left cerebellar artery, Cerebral infarction due to unspecified occlusion or stenosis of unspecified cerebellar artery, Cerebral infarction due to unspecified occlusion or stenosis of other cerebral artery , Cerebral infarction due to cerebral venous thrombosis, nonpyogenic, Other cerebral infarction, Cerebral infarction, unspecified, Occlusion and stenosis of right middle cerebral artery, Occlusion and stenosis of left middle cerebral artery, Occlusion and stenosis of bilateral middle cerebral arteries, Occlusion and stenosis of unspecified middle cerebral artery, Occlusion and stenosis of right anterior cerebral artery, Occlusion and stenosis of left anterior cerebral artery, Occlusion and stenosis of bilateral anterior cerebral arteries, Occlusion and stenosis of unspecified anterior cerebral artery, Occlusion and stenosis of right posterior cerebral artery, Occlusion and stenosis of left posterior cerebral artery, Occlusion and stenosis of bilateral posterior cerebral arteries, Occlusion and |

| Phenotype | Code type | Data codes | Data code definitions |
|---|---|---|---|
| | | I69.922, I69.923, I69.928, I69.931, I69.932, I69.933, I69.934, I69.939, I69.941, I69.942, I69.943, I69.944, I69.949, I69.951, I69.952, I69.953, I69.954, I69.959, I69.961, I69.962, I69.963, I69.964, I69.965, I69.969, I69.990, I69.991, I69.992, I69.993, I69.998, I97.810, I97.811, I97.820, I97.821, Z86.73 | stenosis of unspecified posterior cerebral artery , Occlusion and stenosis of cerebellar arteries, Occlusion and stenosis of other cerebral arteries, Occlusion and stenosis of unspecified cerebral artery, Acute cerebrovascular insufficiency, Cerebral ischemia, Reversible cerebrovascular vasoconstriction syndrome, Other cerebrovascular vasospasm and vasoconstriction , Other cerebrovascular disease, Cerebrovascular Disease, unspecified, Unspecified sequelae of other cerebrovascular disease, Cognitive deficits following other cerebrovascular disease, Aphasia following other cerebrovascular disease, Dysphasia following other cerebrovascular disease, Dysarthria following other cerebrovascular disease, Fluency disorder following other cerebrovascular disease, Other speech and language deficits following other cerebrovascular disease, Monoplegia of upper limb following other cerebrovascular disease affecting right dominant side, Monoplegia of upper limb following other cerebrovascular disease affecting left dominant side, Monoplegia of upper limb following other cerebrovascular disease affecting right non-dominant side, Monoplegia of upper limb following other cerebrovascular disease affecting left non-dominant side, Monoplegia of upper limb following other cerebrovascular disease affecting unspecified side, Monoplegia of lower limb following other cerebrovascular disease affecting right dominant side, Monoplegia of lower limb following other cerebrovascular disease affecting left dominant side, Monoplegia of lower limb following other cerebrovascular disease affecting right non-dominant side, Monoplegia of lower limb following other cerebrovascular disease affecting left non-dominant side, Monoplegia of lower limb following other cerebrovascular disease affecting unspecified side, Hemiplegia and hemiparesis following other cerebrovascular disease affecting right dominant side, Hemiplegia and hemiparesis following other cerebrovascular disease affecting left dominant side, Hemiplegia and hemiparesis following other cerebrovascular disease affecting right non-dominant side, Hemiplegia and hemiparesis following other cerebrovascular disease affecting left non-dominant side, Hemiplegia and hemiparesis following other cerebrovascular disease affecting unspecified side, Other paralytic syndrome following other cerebrovascular disease affecting right dominant side, Other paralytic syndrome following other cerebrovascular disease affecting left dominant side, Other paralytic syndrome following other cerebrovascular disease affecting right non-dominant side, Other paralytic syndrome following other cerebrovascular disease affecting left non-dominant side, Other paralytic syndrome following other cerebrovascular disease, bilateral, Other paralytic syndrome following other cerebrovascular disease affecting unspecified side, Apraxia following other cerebrovascular disease, Dysphagia following other cerebrovascular disease, Facial weakness following other cerebrovascular disease, Ataxia following other cerebrovascular disease, Other sequelae of other cerebrovascular disease , Unspecified Sequelae of unspecified cerebrovascular disease, Cognitive deficits following unspecified cerebrovascular disease, Aphasia following unspecified cerebrovascular disease, Dysphasia following unspecified cerebrovascular disease , Dysarthria following unspecified Cerebrovascular disease, Fluency disorder following unspecified cerebrovascular disease , Other speech and language deficits following unspecified cerebrovascular disease, Monoplegia of upper limb following unspecified cerebrovascular disease affecting right dominant side, Monoplegia of upper limb following unspecified cerebrovascular disease affecting left dominant side, Monoplegia of upper limb following unspecified cerebrovascular disease affecting right non-dominant side, Monoplegia of upper limb following unspecified cerebrovascular disease affecting left non-dominant side, |

| Phenotype | Code type | Data codes | Data code definitions |
|---|---|---|---|
| | | | Monoplegia of upper limb following unspecified cerebrovascular disease affecting unspecified side, Monoplegia of lower limb following unspecified cerebrovascular disease affecting right dominant side, Monoplegia of lower limb following unspecified cerebrovascular disease affecting left dominant side, Monoplegia of lower limb following unspecified cerebrovascular disease affecting right non-dominant side, Monoplegia of lower limb following unspecified cerebrovascular disease affecting left non-dominant side, Monoplegia of lower limb following unspecified cerebrovascular disease affecting unspecified side, Hemiplegia and hemiparesis following unspecified cerebrovascular disease affecting right dominant side , Hemiplegia and hemiparesis following unspecified cerebrovascular disease affecting left dominant side , Hemiplegia and hemiparesis following unspecified cerebrovascular disease affecting right non-dominant side, Hemiplegia and hemiparesis following unspecified cerebrovascular disease affecting left non-dominant side , Hemiplegia and hemiparesis following unspecified cerebrovascular disease affecting unspecified side , Other paralytic syndrome following unspecified cerebrovascular disease affecting right dominant side, Other paralytic syndrome following unspecified cerebrovascular disease affecting left dominant side, Other paralytic syndrome following unspecified cerebrovascular disease affecting right non-dominant side, Other paralytic syndrome following unspecified cerebrovascular disease affecting left non-dominant side, Other paralytic syndrome following unspecified cerebrovascular disease, bilateral, Other paralytic syndrome following unspecified cerebrovascular disease affecting unspecified side, Apraxia following unspecified cerebrovascular disease, Dysphagia following unspecified cerebrovascular disease , Facial weakness following unspecified cerebrovascular disease , Ataxia following unspecified cerebrovascular disease, Other sequelae following unspecified cerebrovascular disease , Intraoperative Cerebrovascular Infarction During cardiac surgery, Intraoperative cerebrovascular infarction during other surgery , Postprocedural cerebrovascular infarction during cardiac surgery, Postprocedural cerebrovascular infarction during other surgery, Personal history of transient ischemic attack (TIA), and cerebral infarction without residual deficits |
| Myocardial Infarction | ICD9 | 410, 410.01, 410.02, 410.1, 410.11, 410.12, 410.2, 410.21, 410.22, 410.3, 410.31, 410.32, 410.4, 410.41, 410.42, 410.5, 410.51, 410.52, 410.6, 410.61, 410.62, 410.7, 410.71, 410.72, 410.8, 410.81, 410.82, 410.9, 410.91, 410.92, 412, 429.79 | Acute myocardial infarction of anterolateral wall, episode of care unspecified, Acute myocardial infarction of anterolateral wall, initial episode of care, Acute myocardial infarction of anterolateral wall, subsequent episode of care , Acute myocardial infarction of other anterior wall, episode of care unspecified, Acute myocardial infarction of other anterior wall, initial episode of care, Acute myocardial infarction of other anterior wall, subsequent episode of care, Acute myocardial infarction of inferolateral wall, episode of care unspecified, Acute myocardial infarction of inferolateral wall, initial episode of care, Acute myocardial infarction of inferolateral wall, subsequent episode of care, Acute myocardial infarction of inferoposterior wall, episode of care unspecified, Acute myocardial infarction of inferoposterior wall, initial episode of care, Acute myocardial infarction of inferoposterior wall, subsequent episode of care, Acute myocardial infarction of other inferior wall, episode of care unspecified, Acute myocardial infarction of other inferior wall, initial episode of care, Acute myocardial infarction of other inferior wall, subsequent episode of care, Acute myocardial infarction of other lateral wall, episode of care unspecified, Acute myocardial infarction of other lateral wall, initial episode of care, Acute myocardial infarction of other lateral wall, subsequent episode of care, True posterior wall infarction, episode of care unspecified, True posterior wall infarction, initial episode of care, True posterior wall infarction, subsequent episode of care, Subendocardial infarction, episode of care unspecified, Subendocardial infarction, initial episode of care, Subendocardial infarction, subsequent |

| Phenotype | Code type | Data codes | Data code definitions |
|---|---|---|---|
| | | | episode of care, Acute myocardial infarction of other specified sites, episode of care unspecified, Acute myocardial infarction of other specified sites, initial episode of care, Acute myocardial infarction of other specified sites, subsequent episode of care, Acute myocardial infarction of unspecified site, episode of care unspecified, Acute myocardial infarction of unspecified site, initial episode of care, Acute myocardial infarction of unspecified site, subsequent episode of care, Old myocardial infarction , Certain sequelae of myocardial infarction, not elsewhere classified, other |
| Myocardial Infarction | ICD10 | I21.01, I21.02, I21.09, I21.11, I21.19, I21.21, I21.29, I21.3, I21.4, I22.0, I22.1, I22.2, I22.8, I22.9, I23.0, I23.1, I23.2, I23.3, I23.4, I23.5, I23.6, I23.7, I23.8, I24.1, I25.2 | ST elevation (STEMI) myocardial infarction involving left main coronary artery, ST elevation (STEMI) myocardial infarction involving left anterior descending coronary artery, ST elevation (STEMI) myocardial infarction involving other coronary artery of anterior wall, ST elevation (STEMI) myocardial infarction involving right coronary artery, ST elevation (STEMI) myocardial infarction involving other coronary artery of inferior wall, ST elevation (STEMI) myocardial infarction involving left circumflex coronary artery, ST elevation (STEMI) myocardial infarction involving other sites, ST elevation (STEMI) myocardial infarction of unspecified site , Non-ST elevation (NSTEMI) myocardial infarction , Subsequent ST elevation (STEMI) myocardial infarction of anterior wall, Subsequent ST elevation (STEMI) myocardial infarction of inferior wall, Subsequent non-ST elevation (NSTEMI) myocardial infarction , Subsequent ST elevation (STEMI) myocardial infarction of other sites, Subsequent ST elevation (STEMI) myocardial infarction of unspecified site, Hemopericardium as current complication following acute myocardial infarction, Atrial septal defect as current complication following acute myocardial infarction, Ventricular septal defect as current complication following acute myocardial infarction, Rupture of cardiac wall without hemopericardium as current complication following acute myocardial infarction, Rupture of chordae tendineae as current complication following acute myocardial infarction, Rupture of papillary muscle as current complication following acute myocardial infarction, Thrombosis of atrium, auricular appendage, and ventricle as current complications following acute myocardial infarction, Postinfarction angina, Other current complications following acute myocardial infarction, Dressler's syndrome, Old myocardial infarction |
| Atrial fibrillation (AF) defined as ECG diagnosis of AF, ≥1 inpatient diagnosis code, ≥1 procedural code, or ≥2 codes of any type[6]<br>Myocardial infarction defined as ≥2 codes from any setting[7]<br>Ischemic stroke defined as ≥2 codes from any setting[8]<br>Heart failure defined as ≥1 inpatient diagnosis code | | | |

**Table 4.** Rule-based approach to train NLP recovery models

| Vital Sign | Context Words | Units Considered | Text Patterns | Labeled Example<br>In format: word {LABEL} |
|---|---|---|---|---|
| Height | “height”,<br>“height:”,<br>“ht”,<br>“ht:” | ‘inches’, ‘in’, ‘feet’, ‘ft’, ‘m’, ‘meters’, ‘cm’, ‘centimeters’, ‘’’ (for feet) ‘”’ (for inches) | [number] | Ht: 63.5 {HEIGHT} |
| | | | [number] [unit] | Patient height is 63.5 {HEIGHT} inches {HEIGHT_UNIT} |
| | | | [number] [unit]<br>[number] [unit] | Height: 5 {HEIGHT} feet {HEIGHT_UNIT} 11 {HEIGHT} inches {HEIGHT_UNIT} |
| Weight | “weight”,<br>“weight:”,<br>“wt”,<br>“wt:” | ‘pounds’, ‘lbs’, ‘lb’, ‘ounces’, ‘oz’, ‘kilograms’, ‘kg, ‘grams’, ‘g’ | [number] | Wt: 180 {WEIGHT} |
| | | | [number] [unit] | Current weight is 65.9 {WEIGHT} kg {WEIGHT_UNIT} |
| | | | [number] [unit]<br>[number] [unit] | Patient’s weight is 170 {WEIGHT} lbs {WEIGHT_UNIT} 9 {WEIGHT} oz {WEIGHT_UNIT} |
| Blood Pressure | “pressure”,<br>“bp”, “bp:” | - | [number]/[number] | Blood pressure is 128/70 {BP} |

**Table 5**. Summary of available data types in C3PO

| Modality | Prior to start of follow-up | | Within 3 years prior to start of follow-up | | During follow-up | | Total (N, % of individuals) | |
|---|---|---|---|---|---|---|---|---|
| | **Total N** | **% of individuals with ≥ 1 study** | **Total N** | **% of individuals with ≥ 1 study** | **Total N** | **% of individuals with ≥ 1 study** | **Total N** | **% of individuals with ≥ 1 study** |
| Electrocardiograms | 954,413 | 40.3 | 471,695 | 28.1 | 1,998,242 | 46.1 | 2,952,655 | 59.1 |
| Echocardiograms | 159,102 | 17.0 | 87,188 | 11.3 | 290,960 | 23.9 | 450,062 | 32.7 |
| Cardiac magnetic resonance imaging | 3,784 | 0.5 | 2,569 | 0.4 | 8,928 | 1.4 | 12,712 | 1.9 |
| Narrative notes | 17,285,956 | 93.9 | 11,766,067 | 92.4 | 59,360,809 | 97.0 | 76,646,765 | 99.5 |
| Head computed tomography | 110,387 | 11.9 | 56,704 | 6.7 | 212,508 | 17.3 | 322,895 | 24.8 |
| Brain magnetic resonance imaging | 86,776 | 8.7 | 47,452 | 5.6 | 170,176 | 15.0 | 256,952 | 20.8 |
| Genetic data (through linkage to MGB Biobank)*† | | | | | | | 42,759 | 8.2 |

*Reported as number of individuals. For all other modalities, reported as number of tests.
†Includes all individuals enrolled in MGB Biobank. Although genotyping is planned in all individuals, it is available in 14,401 (2.8%) as of 03/2021.

**Table 6.** Vital sign-specific yield of NLP recovery

| Vital sign feature | Total NLP values extracted | # with NLP values | # with feature missing in tabular data* | # with ≥ 1 eligible note for feature recovery† | # with recovered feature | Eligible recovery rate (%) |
|---|---|---|---|---|---|---|
| Height | 1,115,248 | 245,217 | 299,905 | 219,873 | 59,682 | 27.1% |
| Weight | 2,749,471 | 310,687 | 204,049 | 143,655 | 76,298 | 53.1% |
| Systolic blood pressure | 3,774,943 | 379,713 | 191,041 | 133,130 | 108,704 | 81.7% |
| Diastolic blood pressure | 1,115,248 | 245,217 | 191,041 | 133,130 | 108,704 | 81.7% |

*Missingness defined as no value included in the tabular data within 3 years prior to start of follow-up
†Eligible notes included discharge summaries, admission notes, and inpatient or outpatient progress notes, within 3 years prior to start of follow-up

**Table 7.** Performance of White PCE models with and without restriction to White individuals

| Model | Hazard ratio (per 1-SD increase) | C-index | GND $\chi^{‡}$ | Recalibrated GND $\chi^{2‡\|\|}$ | ICI[\|\|] | Recalibrated ICI[§\|\|] | Calibration slope[#] |
|---|---|---|---|---|---|---|---|
| *C3PO* | | | | | | | |
| PCE (White women)* | 2.50 (2.41-2.59) | 0.768 (0.759-0.777) | 412, p<0.01 | 1336, p<0.01 | 0.018 (0.016-0.020) | 0.034 (0.032-0.036) | 0.67 (0.65-0.70) |
| PCE (White and non-Black women)* | 2.51 (2.43-2.59) | 0.768 (0.760-0.775) | 487, p<0.01 | 1689, p<0.01 | 0.018 (0.017-0.020) | 0.034 (0.031-0.037) | 0.67 (0.65-0.70) |
| PCE (White men)* | 2.16 (2.09-2.24) | 0.736 (0.727-0.746) | 297, p<0.01 | 504, p<0.01 | 0.024 (0.021-0.027) | 0.032 (0.029-0.035) | 0.71 (0.68-0.74) |
| PCE (White and non-Black men)* | 2.17 (2.11-2.24) | 0.738 (0.730-0.746) | 361, p<0.01 | 618, p<0.01 | 0.024 (0.022-0.027) | 0.032 (0.029-0.035) | 0.70 (0.68-0.73) |

*PCE (White women): 3,442, 90,767, 7.2 (2.8, 10); PCE (White and non-Black women): 4,231, 107,998, 7.1 (2.8, 10); PCE (White men): 4,216, 63,945, 6.4 (2.3, 10); PCE (White and non-Black men): 4,928, 76,304, 6.2 (2.3, 10)†C-index calculated using the inverse probability of censoring weighting method[9]
†C-index calculated using the inverse probability of censoring weighting method[9]
‡Greenwood-Nam-D'Agostino (GND) test, a test of calibration.[10] Lower chi-squared values suggest better calibration (across equally-sized samples). Significant p-values indicate evidence of miscalibration.
§Values after recalibration to the baseline hazard of the sample (see text)
||Integrated calibration index, a quantitative measure of the average difference between predicted event risk and observed event incidence, weighted by the empirical distribution of event risk.[11] Smaller values indicate better calibration.
#A measure of calibration applicable to models that are calibrated-in-the-large.[12,13] A calibration slope equal to one is optimally calibrated.

**Table 8.** Risk model performance in samples with and without NLP recovery

| Model | N events | N total | Hazard ratio (per 1-SD increase) | C-index‡ | GND $\chi^{§}$ | Recalibrated GND $\chi^{2§||}$ | ICI# | Recalibrated ICI#|| | Calibration slope** |
|---|---|---|---|---|---|---|---|---|---|
| *C3PO (without NLP recovery)* | | | | | | | | | |
| PCE (White women)* | 3,110 | 81,870 | 2.51 (2.41-2.60) | 0.768 (0.759-0.776) | 382, p<0.01 | 1319, p<0.01 | 0.019 (0.017-0.021) | 0.036 (0.033-0.039) | 0.66 (0.64-0.69) |
| PCE (Black women)* | 447 | 6,631 | 2.42 (2.15-2.72) | 0.729 (0.703-0.755) | 47, p<0.01 | 185, p<0.01 | 0.029 (0.021-0.037) | 0.056 (0.047-0.065) | 0.61 (0.52-0.69) |
| PCE (White men)* | 3,496 | 56,195 | 2.17 (2.09-2.25) | 0.735 (0.726-0.745) | 280, p<0.01 | 453, p<0.01 | 0.025 (0.022-0.028) | 0.033 (0.030-0.036) | 0.70 (0.66-0.73) |
| PCE (Black men)* | 299 | 4,199 | 2.17 (1.92-2.44) | 0.740 (0.709-0.771) | 23, p<0.01 | 12, p=0.25 | 0.020 (0.0064-0.033) | 0.0041 (0-0.019) | 0.95 (0.81-1.09) |
| CHARGE-AF* | 6,105 | 136,116 | 2.56 (2.50-2.62) | 0.782 (0.776-0.788) | 1366, p<0.01 | 1085, p<0.01 | 0.028 (0.027-0.030) | 0.020 (0.019-0.021) | 0.77 (0.75-0.79) |
| *C3PO (with NLP recovery)* | | | | | | | | | |
| PCE (White women)† | 4,231 | 107,998 | 2.51 (2.43-2.59) | 0.768 (0.760-0.775) | 487, p<0.01 | 1689, p<0.01 | 0.018 (0.017-0.020) | 0.034 (0.031-0.037) | 0.67 (0.65-0.70) |
| PCE (Black women)† | 617 | 8,450 | 2.39 (2.17-2.64) | 0.724 (0.702-0.746) | 69, p<0.01 | 257, p<0.01 | 0.030 (0.023-0.036) | 0.057 (0.050-0.064) | 0.60 (0.53-0.67) |
| PCE (White men)† | 4,928 | 76,304 | 2.17 (2.11-2.24) | 0.738 (0.730-0.746) | 361, p<0.01 | 618, p<0.01 | 0.024 (0.022-0.027) | 0.032 (0.029-0.035) | 0.70 (0.68-0.73) |
| PCE (Black men)† | 425 | 5,432 | 2.04 (1.85-2.25) | 0.725 (0.698-0.751) | 21, p=0.02 | 18, p=0.03 | 0.012 (0-0.025) | 0.010 (0-0.024) | 0.88 (0.77-1.00) |
| CHARGE-AF† | 7,877 | 174,644 | 2.56 (2.50-2.61) | 0.782 (0.777-0.787) | 1856, p<0.01 | 1367, p<0.01 | 0.028 (0.027-0.030) | 0.019 (0.018-0.021) | 0.77 (0.75-0.79) |

*PCE (White women): 6.7 (2.6, 10); PCE (Black women): 6.9 (2.6, 10); PCE (White men): 5.8 (2.2, 10); PCE (Black men): 6.1 (2.2, 10); CHARGE-AF: median follow-up, years (Q1,Q3): 5.0 (2.4,5.0)
†PCE (White women): 7.1 (2.8, 10); PCE (Black women): 7.7 (2.9, 10); PCE (White men): 6.2 (2.3, 10); PCE (Black men): 6.7 (2.5, 10); CHARGE-AF: median follow-up, years (Q1,Q3): 5.0 (2.3,5.0)
‡C-index calculated using the inverse probability of censoring weighting method[9]
§Greenwood-Nam-D'Agostino (GND) test, a test of calibration.[10] Lower chi-squared values suggest better calibration (across equally-sized samples). Significant p-values indicate evidence of miscalibration.
||Values after recalibration to the baseline hazard of the sample (see text)
#Integrated calibration index, a quantitative measure of the average difference between predicted event risk and observed event incidence, weighted by the empirical distribution of event risk.[11] Smaller values indicate better calibration.
**A measure of calibration applicable to models that are calibrated-in-the-large.[12,13] A calibration slope equal to one is optimally calibrated.

**Figure 1.** Longitudinal structure of C3PO

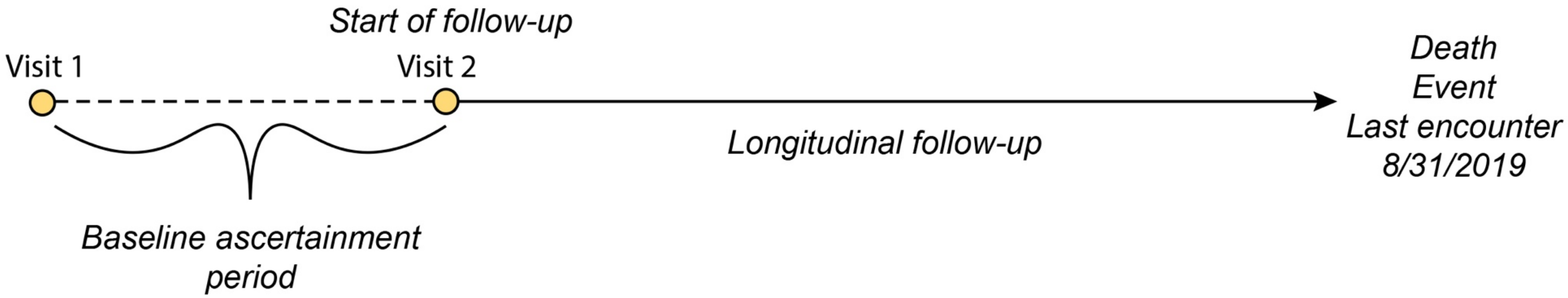

Depicted is an overview of the longitudinal analysis design of C3PO. All individuals meeting inclusion in C3PO had at least one pair of primary care office visits occurring between 1-3 years apart. The start of follow-up for longitudinal analyses for each individual was defined as the second visit of the earliest qualifying pair (i.e., Visit 2 in the diagram). The period between Visit 1 and Visit 2 was included to increase the likelihood that baseline characteristics (e.g., physical measurements, diagnoses) would be appropriately documented in the EHR prior to longitudinal modeling. Longitudinal follow-up then continued until the earliest of death, last encounter in the EHR, or August 31, 2019 (the administrative censoring date of C3PO).

**Figure 2.** Overlap between C3PO and MGH primary care registry

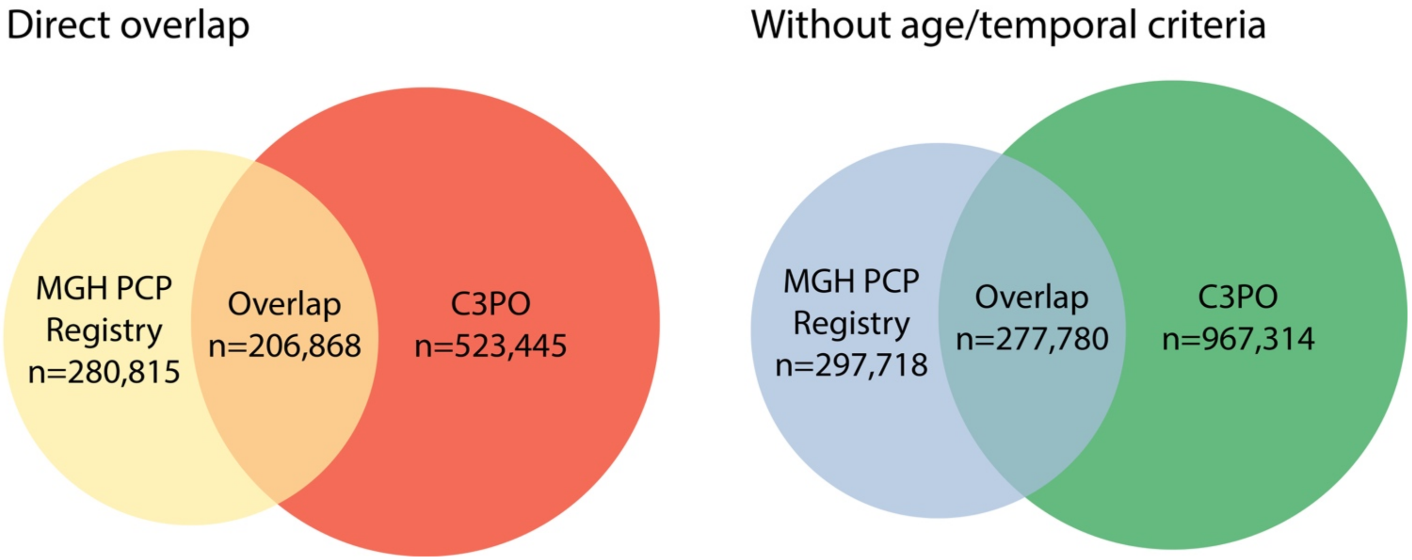

Depicted are Venn diagrams demonstrating overlap between the candidate C3PO cohort and an existing MGH-based primary care registry, to which we applied analogous selection methods. Of 280,815 individuals in the MGH registry meeting the specified temporal selection criteria, the substantial majority (n=206,868; 73.7%) were represented in the candidate C3PO cohort (left panel). The remaining discrepancy was attributed to differences in application of temporal selection criteria (1-3 year windows based on exact dates in C3PO, versus only calendar year data available in MGH registry), as well as exclusion of individuals under age <18 at the start of follow-up in C3PO. Without application of temporal or age selection criteria, 277,780 out of 297,718 (93.3%) of the MGH registry was represented in C3PO (right panel).

**Figure 3**. MI/stroke and AF Convenience Sample construction

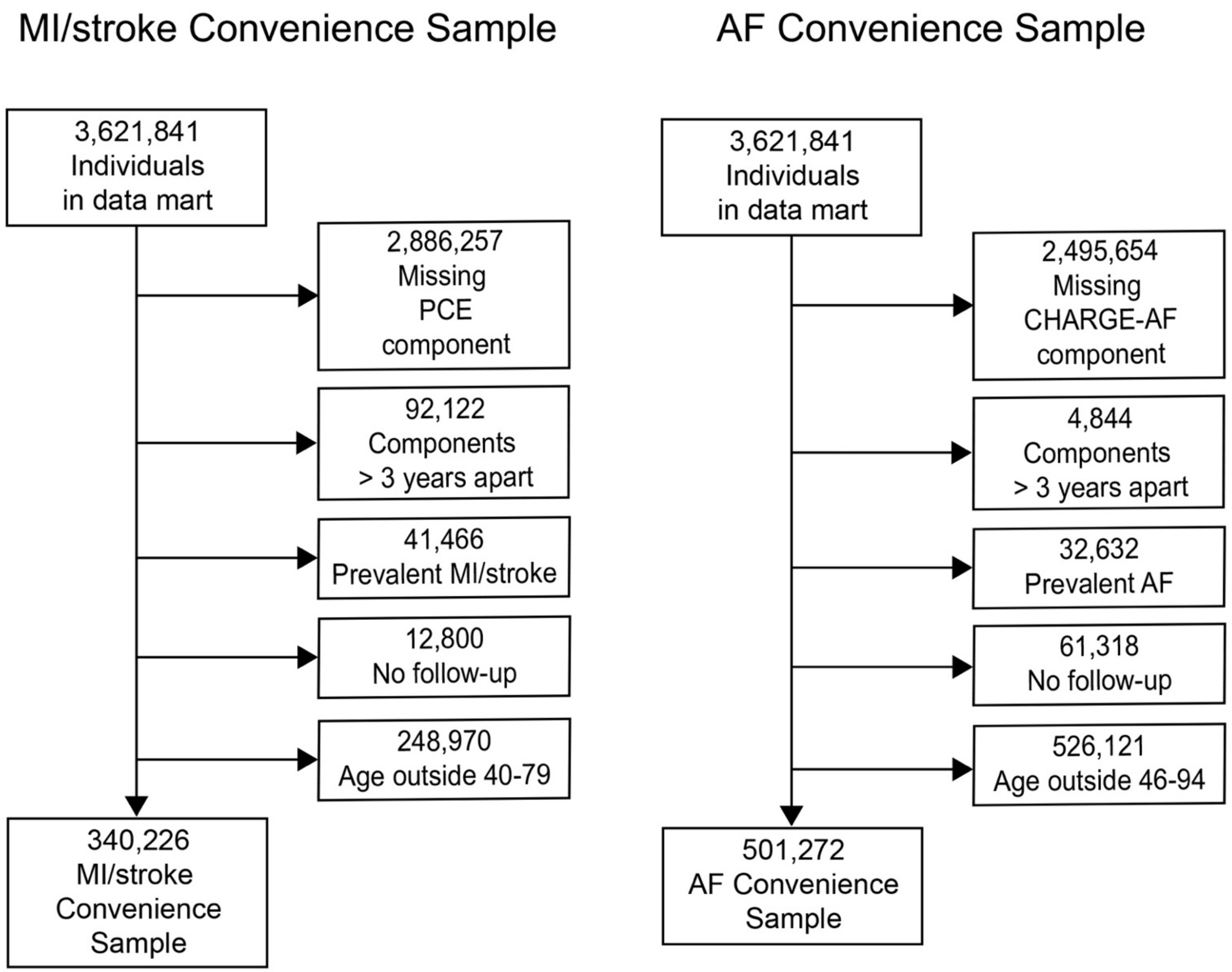

Depicted is an overview of the construction of the AF and MI/stroke Convenience Samples (see **Supplemental Methods 2**).

**Figure 4.** Distribution of office visits and primary care office visits per person in C3PO versus Convenience Samples

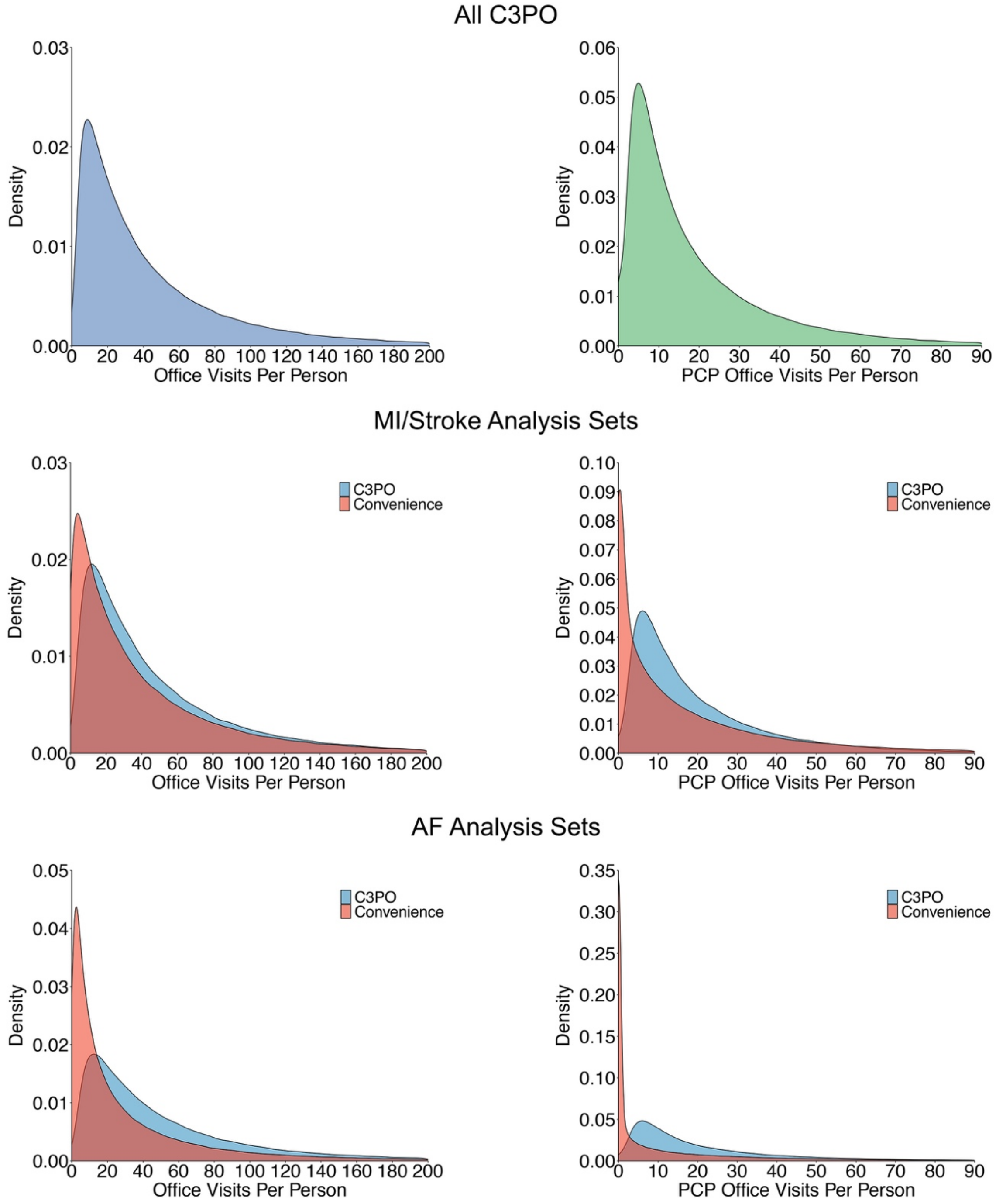

Depicted is the distribution of office visits (left panels) and primary care office visits (right panels) in all of C3PO (top panels), the MI/stroke analysis sets (middle panels), and the AF analysis sets (bottom panels). The MI/stroke and AF analysis sets depict distributions both for C3PO (blue) and the respective Convenience Samples (red).

**Figure 5.** Flow diagrams for C3PO PCE and CHARGE-AF samples

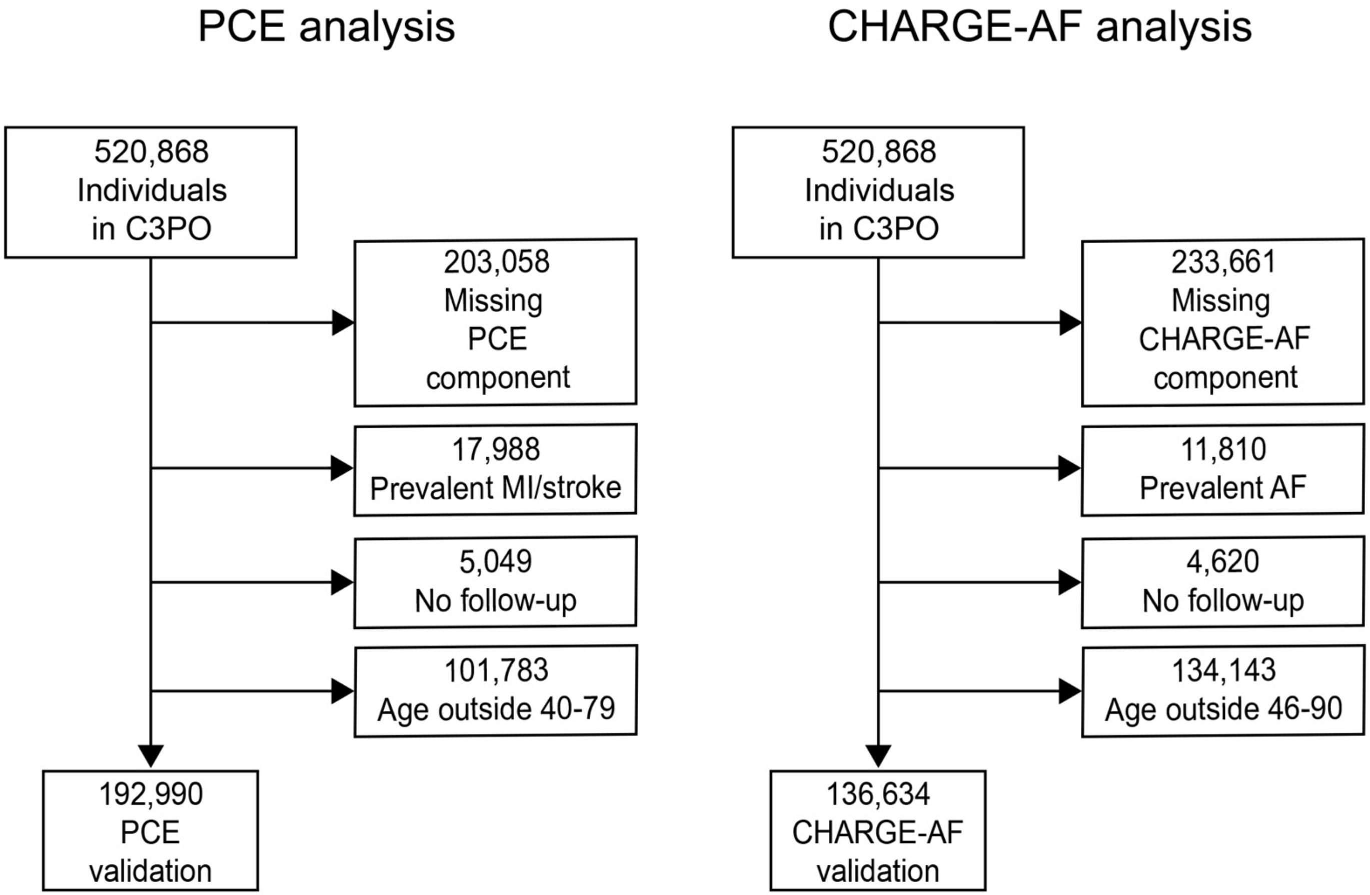

Depicted is the construction of the C3PO-based CHARGE-AF and PCE analysis samples, including relevant exclusions.

**Figure 6.** Predicted event risk in C3PO CHARGE-AF and PCE analysis sets

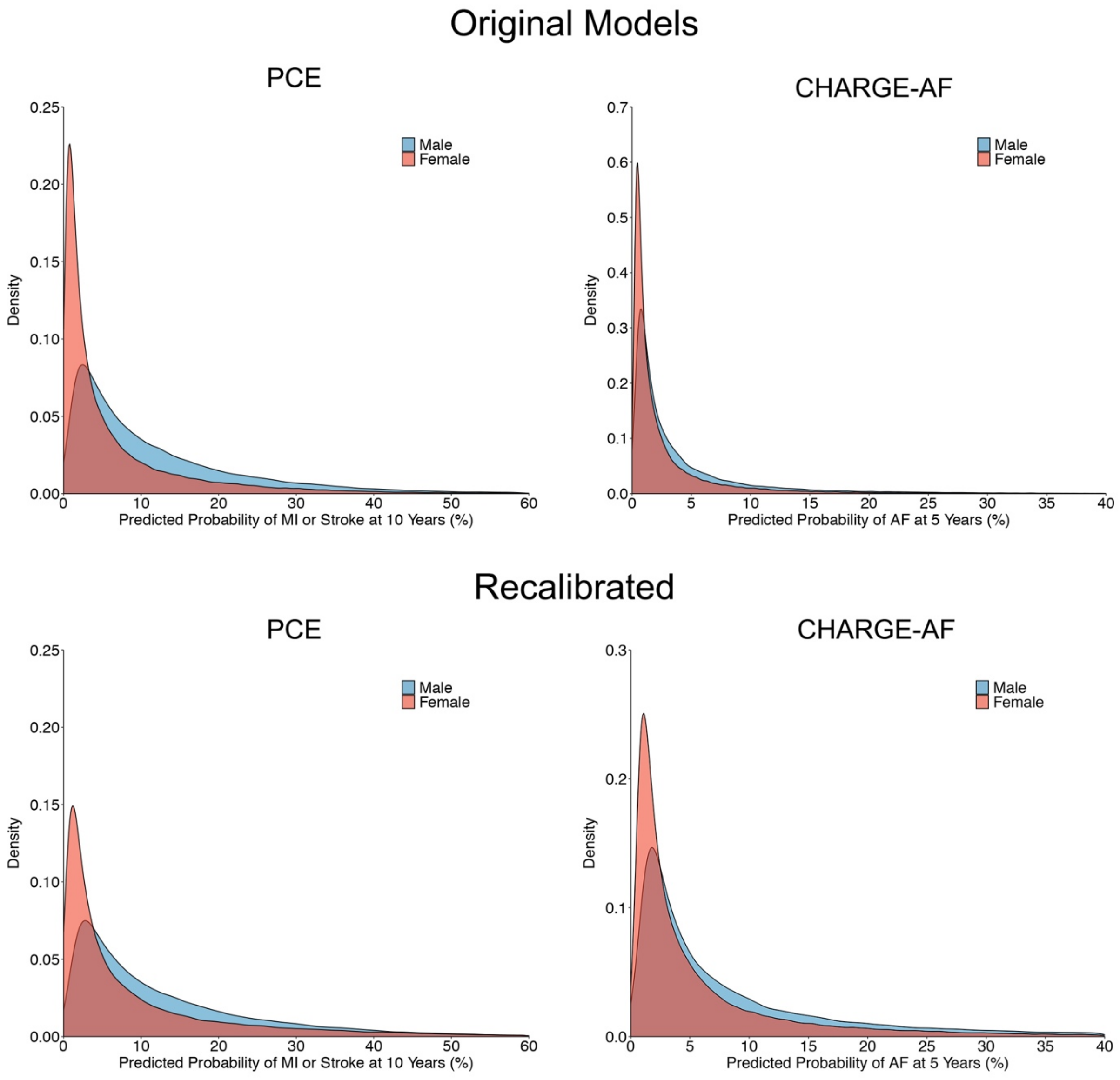

Depicted is sex-stratified the distribution of predicted 10-year MI/stroke risk according to PCE models (left panels) and the 5-year predicted AF risk according to the CHARGE-AF score (right panels). Females are depicted in red and males in blue. Top panels reflect predicted risks obtained using the original published equations. Bottom panels reflect predicted risks after recalibration to the C3PO baseline hazard.

**Figure 7.** Cumulative risk of events stratified by predicted risk

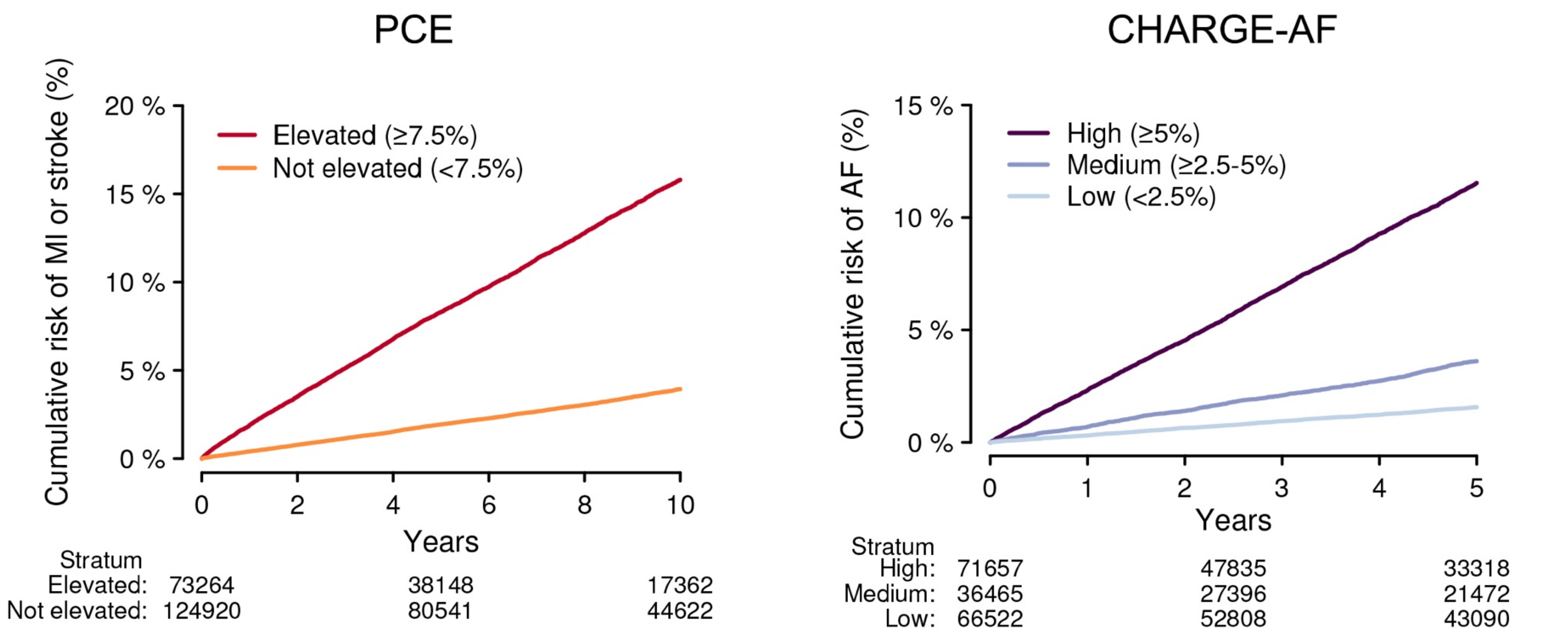

Depicted are the cumulative risks MI/stroke (left) and AF (right), stratified by level of predicted risk according to the Pooled Cohort Equations (PCE, left) and the CHARGE-AF score (right). Given substantial miscalibration using the original CHARGE-AF score, the model recalibrated to the baseline AF hazard of the sample was used for stratification. The risk levels depicted are those defining elevated atherosclerotic disease risk in the 2019 American Heart Association/American College of Cardiology primary prevention guidelines[14] (Elevated: ≥7.5%, Not elevated: <7.5%), and those previously proposed in the original CHARGE-AF derivation study[15] (High: ≥7.5%, Medium: ≥2.5-5%, Low <2.5%), respectively. The number of individuals remaining at risk is depicted below each plot. The x-axis depicts the number of years since start of follow-up, corresponding to the relevant prediction window for each score.

**Figure 8.** Calibration of original models in C3PO and Convenience Samples

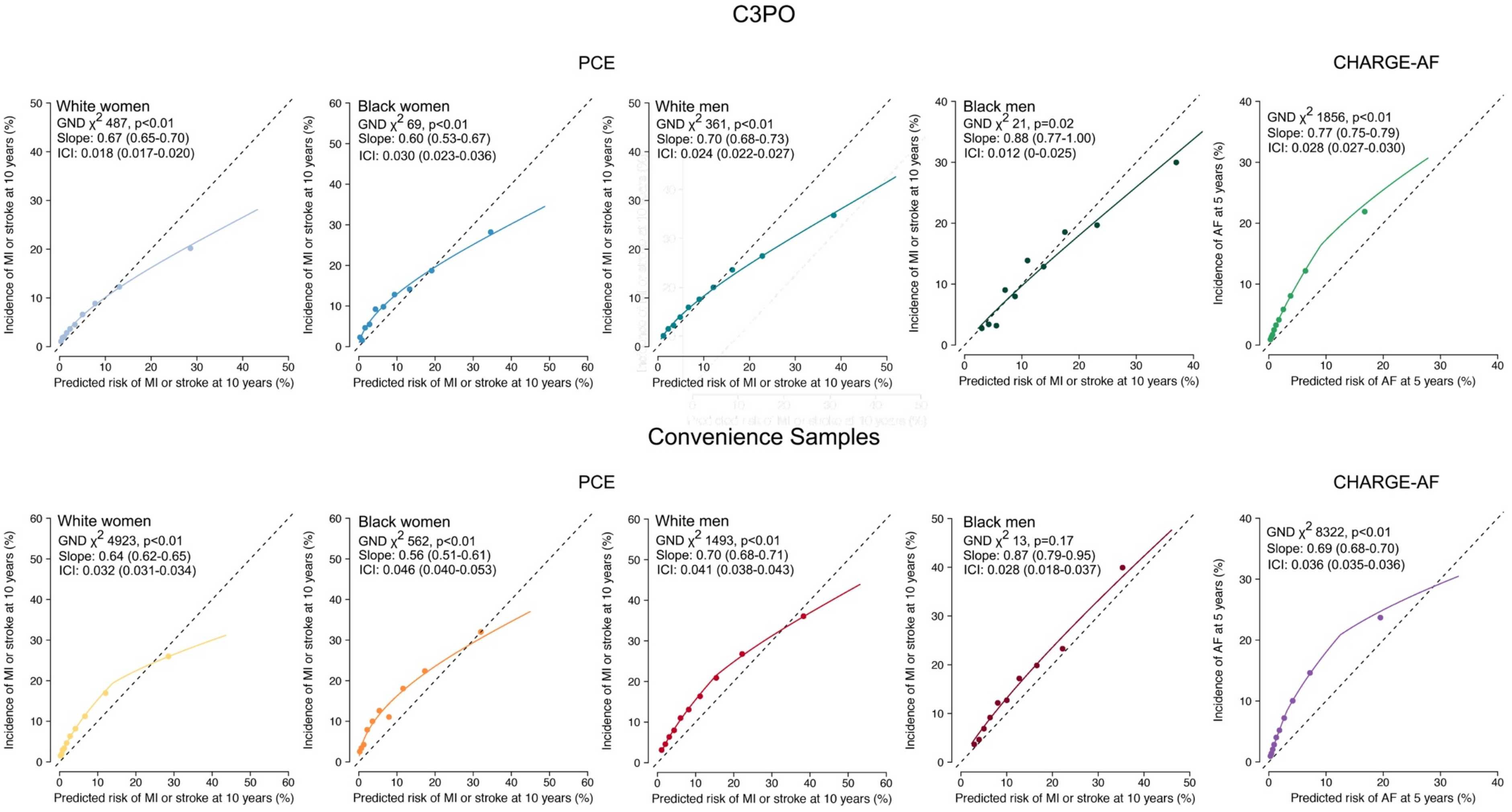

Depicted are calibration plots demonstrating agreement between predicted event risk (x-axis) and observed cumulative event incidence (y-axis) for the Pooled Cohort Equations (PCE, left four panels) and the CHARGE-AF score (right panels) in C3PO (upper panels) versus the Convenience Samples (bottom panels). In each plot, each point represents a decile of predicted risk. Events correspond to the prediction target of each score (i.e., 10-year MI or stroke for the PCE and 5-year incident AF for CHARGE-AF). Perfect calibration is indicated by the hashed diagonal line, denoting perfect correspondence between predicted and observed risk. The Greenwood-Nam-D'Agostino test of calibration is shown for each plot (where a lower chi-squared value indicates better fit), the calibration slope (where a calibration slope of one is optimal), and integrated calibration index (ICI, a measure of prediction error in which lower values indicate more accurate predictions) is shown for each model. Since the PCE score comprises four separate models stratified on the basis of sex and race, the curve for each score is represented separately (see legend). Each plot also depicts a fitted calibration curve obtained using adaptive hazard regression[11] relating predicted risk and observed event risk.

**Figure 9.** Calibration of recalibrated models in C3PO and Convenience Samples

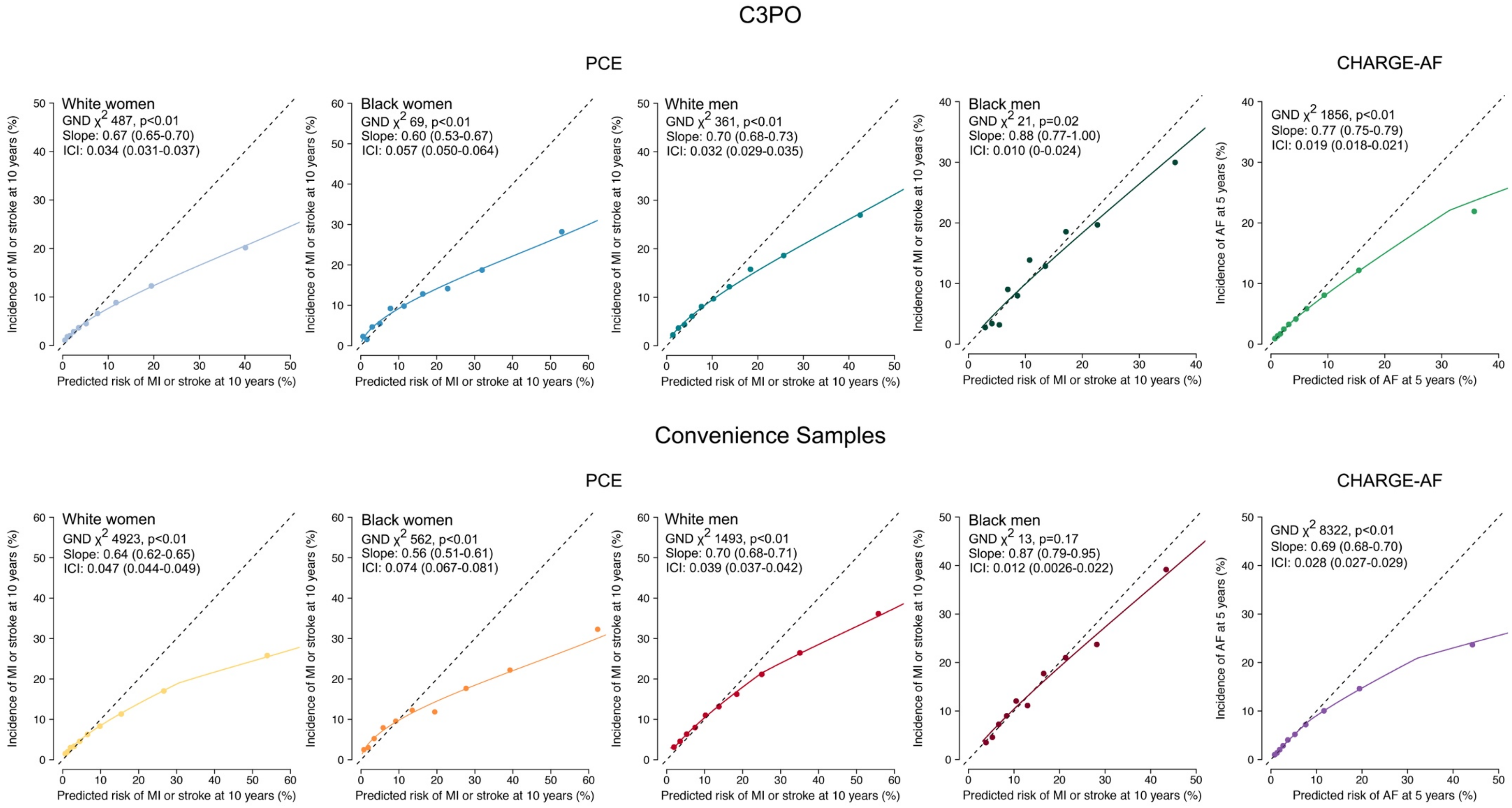

Depicted are calibration plots demonstrating agreement between predicted event risk (x-axis) and observed cumulative event incidence (y-axis) for the Pooled Cohort Equations (PCE, left four panels) and the CHARGE-AF score (right panels) in C3PO (upper panels) versus the Convenience Samples (bottom panels). In each plot, each point represents a decile of predicted risk. Events correspond to the prediction target of each score (i.e., 10-year MI or stroke for the PCE and 5-year incident AF for CHARGE-AF). Perfect calibration is indicated by the hashed diagonal line, denoting perfect correspondence between predicted and observed risk. The Greenwood-Nam-D'Agostino test of calibration is shown for each plot (where a lower chi-squared value indicates better fit), the calibration slope (where a calibration slope of one is optimal), and integrated calibration index (ICI, a measure of prediction error in which lower values indicate more accurate predictions) is shown for each model. Since the PCE score comprises four separate models stratified on the basis of sex and race, the curve for each score is represented separately (see legend). Each plot also depicts a fitted calibration curve obtained using adaptive hazard regression[11] relating predicted risk and observed event risk.
